# Genetic prediction of long-term effects of aromatase inhibition on cancer and non-neoplastic disease risk

**DOI:** 10.64898/2026.04.28.26351848

**Authors:** Devleena Ray, Tessa Bate, Tracy O’Mara, Peter Sasieni, Marc J Gunter, Richard M Martin, Karl Smith-Byrne, Philip C Haycock, James Yarmolinsky

**Affiliations:** MRC Integrative Epidemiology Unit, University of Bristol, Bristol, United Kingdom; Department of Genetics and Computational Biology, QIMR Berghofer Medical Research Institute, Brisbane, Australia; Wolfson Institute of Population Health, Faculty or Medicine & Dentistry, Queen Mary University of London; Department of Epidemiology and Biostatistics, School of Public Health, Imperial College London, London, United Kingdom; Cancer Epidemiology Unit, Oxford Population Health, University of Oxford, Oxford, United Kingdom

## Abstract

**Background:** Anastrozole, an aromatase inhibitor, is approved for breast cancer prevention in high-risk women. The long-term effects of aromatase inhibition, including its repurposing potential to other cancers, possible adverse effects, and treatment effect heterogeneity across patient subgroups, remain unclear.

**Methods:** We used the rs727479 variant in *CYP19A1* to mimic the effect of long-term pharmacological aromatase inhibition. To evaluate repurposing opportunities, genetic association data on five cancers (211,386 cases, 684,665 controls) were obtained from genome-wide association study consortia. Potential adverse effects were evaluated in a phenome-wide association study (PheWAS) of 449 health-related traits in 162,360 postmenopausal women in the UK Biobank. Effects were investigated across clinically relevant subgroups in the UK Biobank including those defined by body mass index (BMI).

**Results:** Genetically-proxied aromatase inhibition was associated with reduced risk of ER+ breast cancer (OR:0·78, 95%CI:0·67-0·92) and decreased heel bone mineral density (−0·32SD change, 95%CI:-0·36,-0·28). When examining the repurposing potential of anastrozole to other cancers, we found that genetically-proxied aromatase inhibition reduced endometrial cancer risk (OR:0·34, 95%CI:0·26-0·44). In PheWAS, genetically-proxied aromatase inhibition was associated with 6 outcomes (*P*_FDR_<0·05) including reduced risk of endometrial polyps (OR:0·58, 95%CI:0·45-0·74) and postmenopausal bleeding (OR:0·67, 95%CI:0·54-0·83), with stronger effects in women with higher BMI (*P*_LRT_=1·26×10^-3^ and 0·02, respectively).

**Conclusion:** Our genetic analyses recapitulate known effects of aromatase inhibition on breast cancer risk and highlight potential repurposing for endometrial cancer prevention. Limited evidence of adverse effects beyond bone mineral density was observed, and subgroup analyses suggested that women with higher BMI may experience greater protection against endometrial conditions.

## Introduction

Breast cancer is the most common form of cancer in women worldwide, with over 2 million cases diagnosed in 2022.^1^ Most invasive breast cancers are hormone-receptor positive (HR+) and driven by oestrogen signalling.^2,3^ Anastrozole, an aromatase inhibitor that reduces oestrogen synthesis, is a standard adjuvant therapy for postmenopausal women with early-stage HR+ breast cancer.^4^

Given the importance of oestrogen in breast carcinogenesis, the International Breast Cancer Intervention Study (IBIS)-II trial evaluated the effect of 5-year anastrozole treatment for breast cancer prevention in high-risk postmenopausal women. The trial randomised 3,864 women and found a 53% reduction in breast cancer incidence compared with placebo (HR:0·47, 95% CI:0·32-0·68) that persisted through 12 years of follow-up with no excess risk of serious adverse events.^5,6^ Based on these findings, the UK Medicine and Health Regulatory Authority (MHRA) approved anastrozole for breast cancer prevention in high-risk postmenopausal women in 2023.^7^

However, several unresolved questions remain regarding the use of anastrozole for breast cancer prevention. First, the patient subgroups most likely to benefit from this medication are unclear. Though exploratory analyses in IBIS-II found no heterogeneity by baseline characteristics, a recent re-analysis reported greater benefit among women with a higher oestradiol-sex hormone-binding globulin (SHBG) ratio and, conversely, little to no benefit among those with lower ratios.^8^ These findings suggest that preventive aromatase inhibition could potentially be targeted to women most likely to respond to endocrine therapy, but require further validation. Second, IBIS-II did not demonstrate a clear reduction in endometrial cancer risk, despite the central role of oestradiol in endometrial carcinogenesis and the substantial risk reduction observed with anastrozole compared to tamoxifen in the Arimidex, Tamoxifen, Alone or in Combination (ATAC) trial.^9^ Whether this discrepancy reflects limited statistical power in IBIS-II (i.e. 12 endometrial cancer cases were diagnosed across study arms) or the choice of tamoxifen as comparator in the ATAC trial is unclear and thus whether the preventive effects of anastrozole extend to other cancers has not been established. Finally, though IBIS-II did not detect increased rates of serious adverse events at 12 years follow-up, higher rates of less serious side effects (e.g. hypertension, arthralgia, carpal tunnel syndrome, infections) were observed during the 5-year active therapy period, which can affect patients’ quality of life and thus treatment adherence.^10^ These adverse events were not measured after completion of active therapy and therefore whether anastrozole has long-term effects on these and other adverse events is not known.

In the absence of long-term clinical trial data, human genetics can provide complementary insight into the on-target effects of approved medications. Specifically, genetic variants located in or near genes encoding targets of approved medications can be used to mimic pharmacological perturbation of these drug targets.^11^ This approach leverages the natural randomisation of germline genetic variants at meiosis to minimise confounding and reverse causation and has been used successfully previously to anticipate clinical benefits and adverse effects of therapeutic interventions.^11–15^

Here, we used the rs727479 variant in *CYP19A1*, encoding aromatase, to mimic the long-term effect of pharmacological aromatase inhibition. We validated rs727479 as a genetic instrument by showing concordant effects of genetically-proxied aromatase inhibition with the effects of anastrozole on disease outcomes and traits reported in IBIS-II. We then investigated the repurposing potential of this medication to other cancers by using data from large-scale cancer genome-wide association studies (GWAS). To identify possible adverse effects beyond those measured in IBIS-II, we performed a phenome-wide association study (PheWAS) of 449 disease and health-related outcomes in 162,360 postmenopausal women in the UK Biobank. Finally, to identify patient subgroups most likely to benefit from anastrozole, we examined the effect heterogeneity by body mass index (BMI), testosterone-to-SHBG ratio, polygenic risk score (PRS) for breast cancer and PRS for bone mineral density (BMD) in the UK Biobank.

## Methods

A schematic overview of the analytical stages of this study is presented in **Figure 1**.

**Figure 1.**
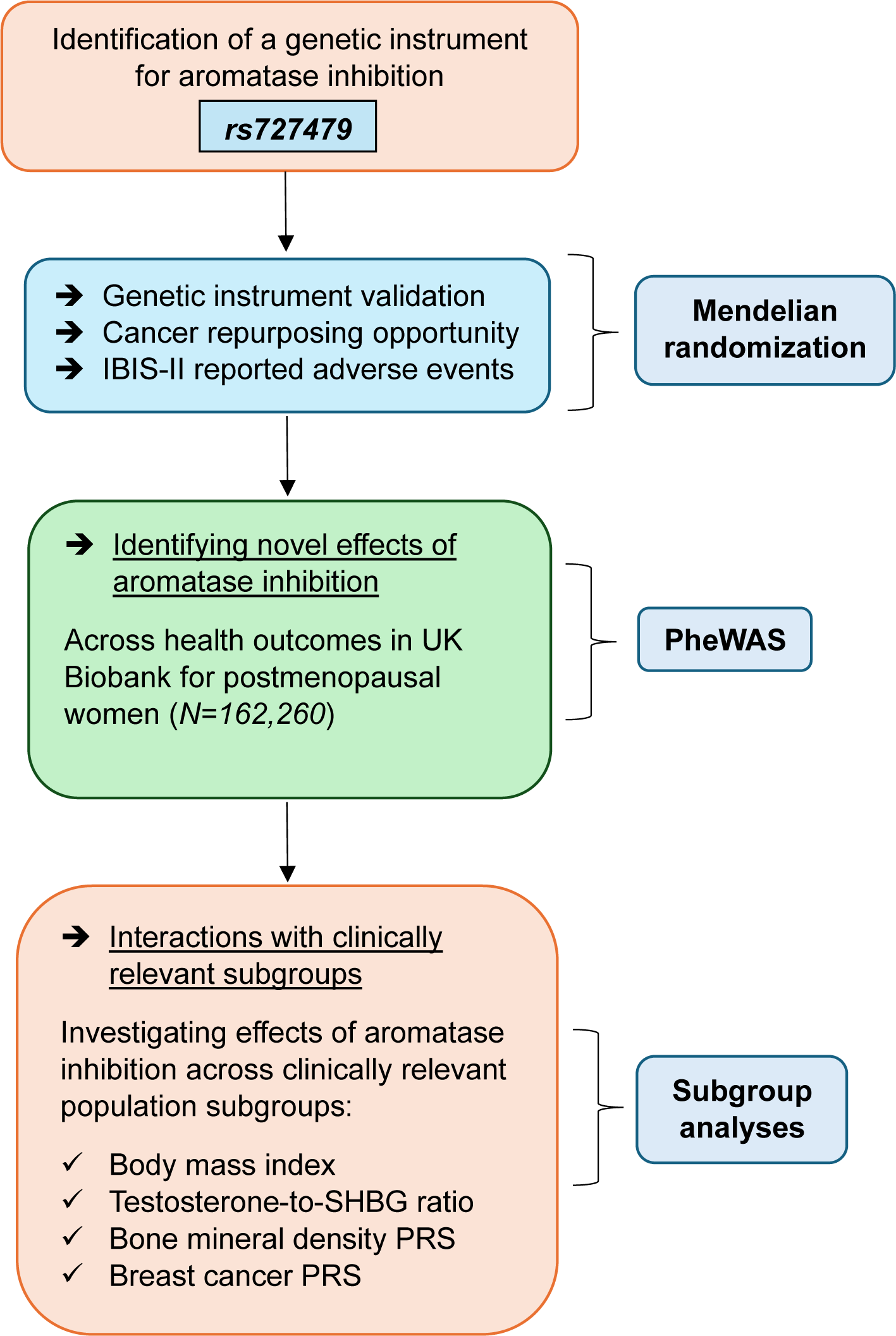
Methods overview

### Genetic instrument selection and validation

We used rs727479, an intronic variant within the *CYP19A1* gene that encodes the aromatase enzyme, as a genetic instrument for aromatase inhibition.^16^ The minor allele (C) of this variant has previously been shown to associate with log-transformed serum oestradiol levels (−0·096, 95% CI:-0·131 to −0·061, *P* = 7·40 x 10^-8^) in 2,767 post-menopausal women of European ancestry, thus mimicking pharmacological aromatase inhibition of oestradiol levels. Using existing methods,^17^ we estimated the proportion of variance in serum oestradiol explained by our instrument (R^2^ statistic) as well as the strength of our instrument (F-statistic). Our genetic instrument explained 1·01% of the variance in serum oestradiol and had an F-statistic of 28·4, suggesting a low likelihood of weak instrument bias in subsequent genetic analyses.

To assess the validity of rs727479 as a genetic instrument, we evaluated the effect of this variant on risk of overall breast cancer, oestrogen receptor-positive (ER+) breast cancer, oestrogen receptor-negative (ER-) breast cancer and bone mineral density (BMD). Summary genetic association data on breast cancer and its subtypes were obtained from a GWAS of breast cancer susceptibility in 122,977 cases and 105,974 controls of European ancestry.^18^ Summary data for heel BMD were obtained from a GWAS in 583,314 individuals of European ancestry.^19^

### Evaluation of repurposing potential of anastrozole to other cancers

To evaluate the possibility of repurposing anastrozole to other cancers, we examined the effect of genetically-proxied aromatase inhibition on the risk of five additional cancers that were evaluated in IBIS-II: colorectal, endometrial, lung, skin, and ovarian cancer. We obtained summary genetic association data on susceptibility to these cancers from previously published GWAS performed in individuals of European ancestry (number of cases across GWAS ranged from 12,906 to 78,473). (**Table 1**)

### Examination of IBIS-II reported adverse effects

To assess if IBIS-II reported adverse effects of anastrozole, observed during the 5-year active therapy period (i.e. hypertension, arthralgia, carpal tunnel syndrome, and influenza), are sustained in the long-term, we evaluated the effect of genetically-proxied aromatase inhibition on these outcomes. Details on the GWAS used to evaluate each outcome are presented in **Table 1** and further information on statistical analysis, imputation and quality control measures adopted by these studies are available in the original publications.^20–26^ We could not evaluate five additional adverse events reported in IBIS-II (i.e. joint stiffness, vasomotor symptoms, dry eyes, vaginal dryness, otitis media) because of the absence of genetic association data on these outcomes.

### Identification of potential novel adverse effects of anastrozole

To identify possible adverse effects of anastrozole beyond those measured in IBIS-II, we performed a PheWAS examining the effect of genetically-proxied aromatase inhibition on 449 disease and health-related outcomes in the UK Biobank using PHESANT.^27^ The analysis was restricted to 162,360 post-menopausal women and adjusted for age at recruitment and the first 10 principal components (PCs) of genetic ancestry. Further details on dataset preparation and the PheWAS analysis are provided in the **Supplementary Methods**.

### Interactions with clinically relevant subgroups

Exploratory subgroup analyses within the IBIS-II trial did not find evidence for heterogeneity of the effect of anastrozole on invasive breast cancer risk by age, body mass index (BMI), HER2 status, or benign breast disease. However, a subsequent re-analysis reported greater benefit among women with medium to higher baseline oestradiol-SHBG ratios. To validate these findings and extend analyses to outcomes not assessed in IBIS-II, we conducted subgroup analyses of genetically proxied aromatase inhibition across strata defined by BMI, testosterone-to-SHBG ratio, breast cancer PRS and bone mineral density PRS in the UK Biobank.

Subgroup analyses were performed in postmenopausal women for outcomes associated with genetically-proxied aromatase inhibition (*P*<0·05) and for outcomes identified in the PheWAS at *P*_FDR_<0·05. Participants were stratified into tertiles of each subgroup variable and Cox proportional hazards models or linear models, as appropriate, were used to estimate the effect on outcomes. ^24^ Models were adjusted for age at recruitment and the first ten principal components of genetic ancestry. Statistical evidence for an interaction was assessed using the log-likelihood ratio test (LRT) comparing models with and without an interaction term between rs727479 and a continuous measure of the subgroup variable. The rationale for subgroup selection and further details on the statistical analyses are provided in the **Supplementary Methods**.

## Results

### Genetic instrument validation

Genetically-proxied aromatase inhibition was associated with a reduced risk of overall breast cancer (OR:0·85, 95% CI:0·74-0·97) and ER+ breast cancer (OR:0·78, 95% CI:0·67-0·92), and a decrease in heel BMD (−0·32 SD units, 95% CI:-0·36 to −0·28) per SD reduction in oestradiol (**Figure 2; Supplementary Table 1**). There was little evidence of an association with risk of ER-breast cancer (OR:0·94, 95% CI:0·74-1·20), compatible with findings from IBIS-II. These findings are consistent with the effects of anastrozole in the IBIS-II trial, thus validating our instrument as a suitable proxy for aromatase inhibition.

**Figure 2.**
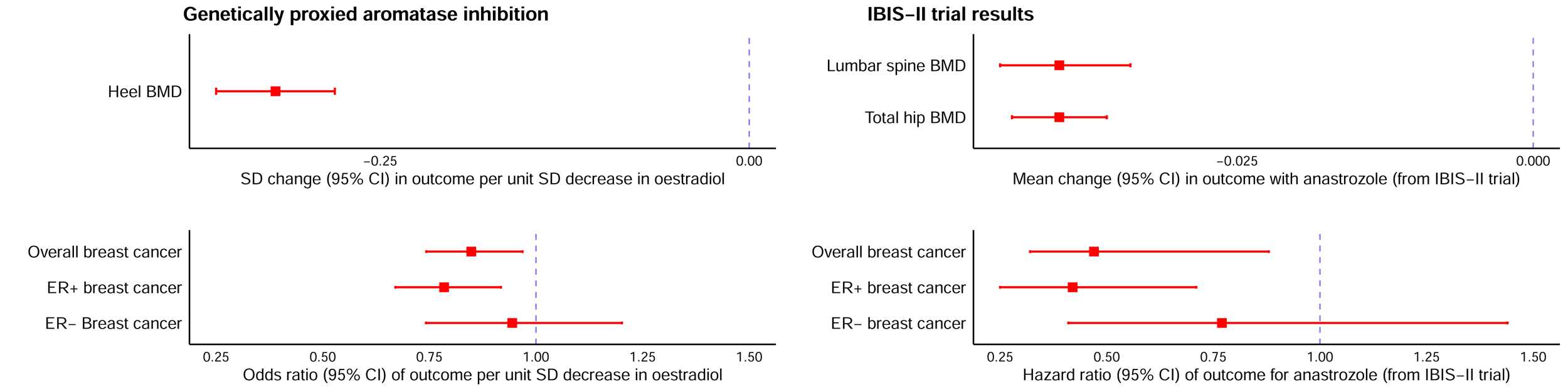
Comparison of genetically proxied aromatase inhibition and IBIS-II estimates for established effects of anastrozole. IBIS-II estimates were obtained from the published trial report.^6^

### Evaluation of repurposing potential of anastrozole to other cancers

We found strong evidence that genetically-proxied aromatase inhibition lowered risk of endometrial cancer (OR 0·34, 95% CI 0·26-0·44) (**Figure 3; Supplementary Table 1**). Stratification by histological subtype revealed a stronger protective association for endometrioid endometrial cancer (OR:0·26, 95% CI:0·18-0·39), than non-endometrioid cancer (OR:0·71, 95% CI:0·28-1·83). Though directionally consistent with our findings, the IBIS-II trial found limited statistical evidence of an effect of anastrozole on endometrial cancer (HR:0·72, 95% CI:0·18-2·65) which could reflect low statistical power due to the small number of incident cancers observed in that study (7 vs 10 in anastrozole and placebo arms, respectively). We found some evidence that genetically-proxied aromatase inhibition lowered risk of non-melanoma skin cancer (OR:0·83, 95% CI:0·68-1·01) which was concordant with reduced risk of this cancer reported in IBIS-II (HR:0·59, 95% CI:0·39-0·87). We found little evidence of an effect of genetically-proxied aromatase inhibition on the risk of colorectal, lung, and ovarian cancer, consistent with findings from IBIS-II.

**Figure 3.**
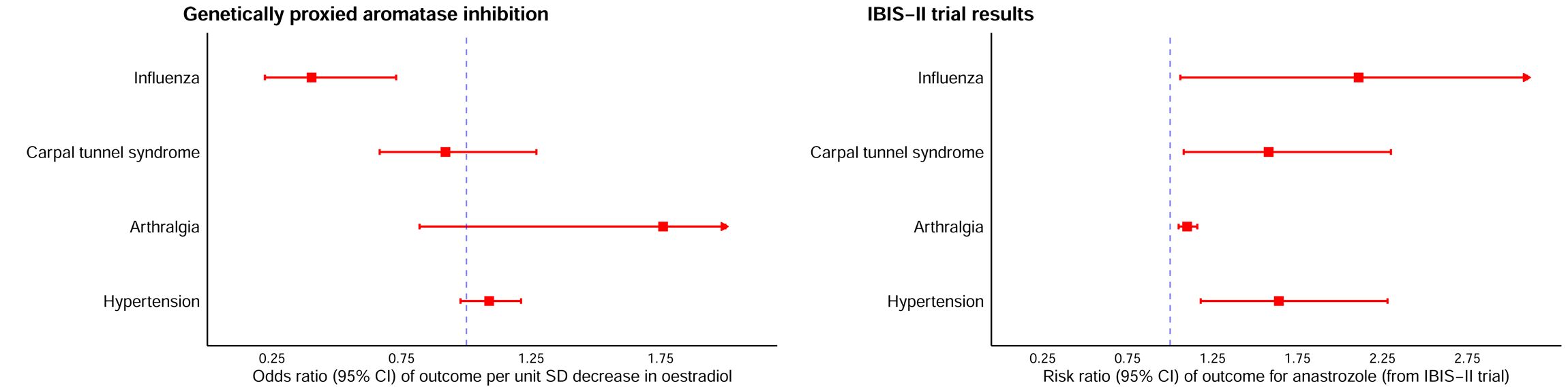
Comparison of genetically proxied aromatase inhibition and IBIS-II estimates for cancer outcomes. *Malignant non-melanoma skin cancer was assessed in our study whereas IBIS-II had no information on malignancy of non-melanoma skin cancer that was reported in the study. IBIS-II estimates were obtained from the published trial report.^6^

### Examination of IBIS-II reported adverse effects

We found weak evidence that genetically-proxied aromatase inhibition increased long-term hypertension risk (OR:1·08, 95% CI:0·98-1·21) which was concordant in direction with the IBIS-II reported increased risk of hypertension during the 5-year active therapy period (**Figure 4; Supplementary Table 1**). While IBIS-II reported increased rates of arthralgia and carpal tunnel syndrome during active therapy, our long-term genetic estimates were imprecise (OR:1·75, 95% CI:0·82-3·78, and OR:0·92, 95% CI:0·66-1·27, respectively), and thus possible effects on these outcomes cannot be ruled out. Contrary to an increased risk of influenza reported in IBIS-II, we found evidence that genetically-proxied aromatase inhibition lowered long-term influenza risk (OR:0·40, 95% CI:0·22-0·73).

**Figure 4.**
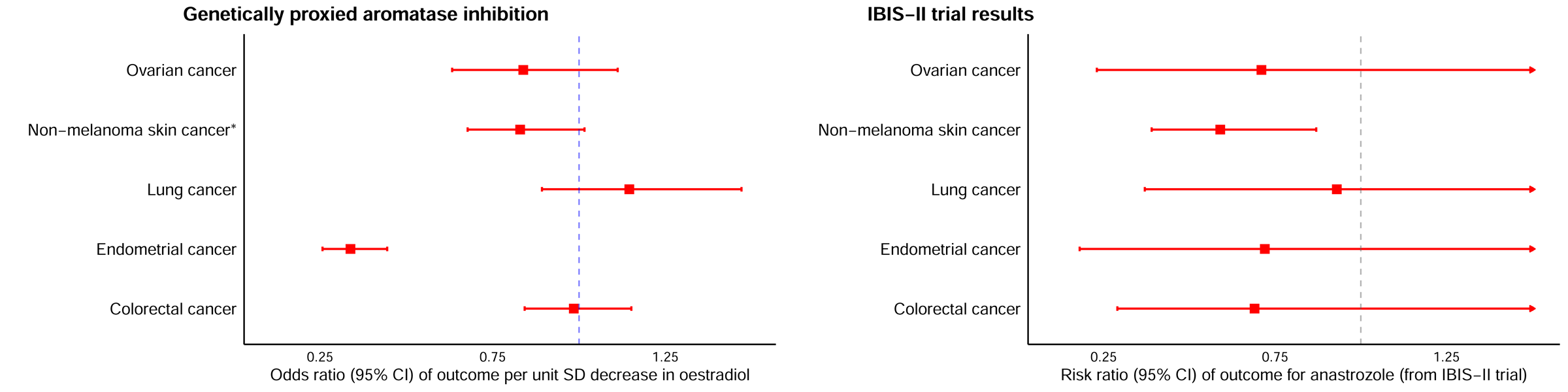
Comparison of genetically proxied aromatase inhibition and IBIS-II estimates* for adverse secondary effects of anastrozole. *The estimates for the IBIS-II reported adverse events were obtained from the original 5-year trial data as these outcomes were not included in the long follow-up (12-year) study.^5^

### Identification of potential novel adverse effects of anastrozole

In a PheWAS of 449 disease and health-related outcomes in the UK Biobank that were not evaluated in IBIS-II, we identified 6 traits strongly associated with genetically-proxied aromatase inhibition after accounting for multiple testing (*P*_FDR_<0·05). 5 of these traits showed protective associations, including postmenopausal bleeding (OR:0·67, 95% CI:0·54-0·83), endometrial polyps (OR:0·58, 95% CI:0·45-0·74), malaise and fatigue (OR:0·49, 95% CI:0·33-0·72), infection following a procedure (OR:0·46, 95% CI:0·29-0·70) and cancer chemotherapy session (OR:0·59, 95% CI:0·45-0·77). In contrast, genetically-proxied aromatase inhibition was positively associated with distal radius fracture risk (OR:1·92, 95% CI:1·39-2·66). Complete findings from PheWAS analyses are presented in **Figure 5** and **Supplementary Table 2.**

**Figure 5.**
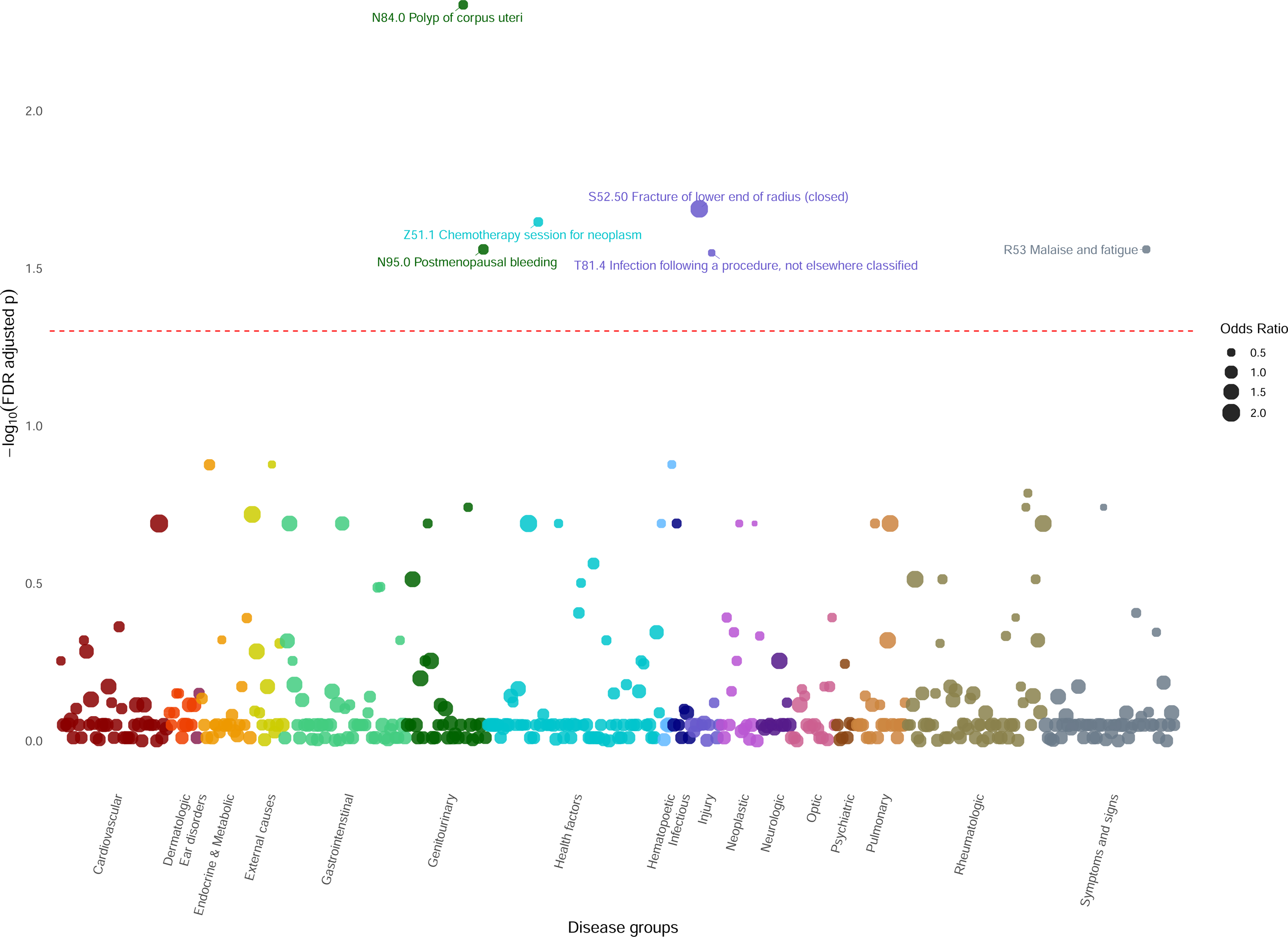
Phenome-wide association study (PheWAS) Manhattan Plot. Each point on the plot represents the association between genetically-proxied aromatase inhibition and a trait. The point size reflects the magnitude of effect and the colours indicate outcome categories, grouped according to the ICD-10 based classifications used in UK Biobank. The dashed horizontal line marks the FDR-adjusted threshold and the most significant phenotype-SNP associations across all health outcomes are text-labelled. For comparability with our MR findings, the effect estimates were converted into MR-scaled odds ratio by calculating a Wald ratio and corresponding standard error (SE) derived using the delta method.

### Interactions with clinically relevant subgroups

In subgroup analyses, we found evidence of interaction between BMI and the effect of genetically-proxied aromatase inhibition on risk of endometrial polyps and postmenopausal bleeding (**Figure 6**). Specifically, we found evidence that genetically-proxied aromatase inhibition was associated with more pronounced reductions in risk of endometrial polyps in women in the upper (HR:0·87, 95% CI:0·82-0·92) and middle tertiles (HR:0·93, 95% CI:0·88-0·98) of BMI but not in the lowest BMI tertile (HR:1·02, 95% CI:0·97-1·08; *P*_LRT_=1·26×10^−3^). Genetically-proxied aromatase inhibition was also associated with reduced risk of postmenopausal bleeding in women in the upper BMI tertile (HR:0·89, 95% CI:0·85-0·94) but not in the middle (HR:0·96, 95%:CI 0·92-1·01) or lower tertiles (HR:0·97, 95% CI:0·93-1,02; *P*_LRT_=0·02). We found little-to-no evidence of interaction across subgroup analyses of testosterone-to-SHBG ratio or polygenic risk scores for breast cancer or BMD after accounting for multiple testing. Complete findings from subgroup analyses are presented in **Supplementary Table 3**.

**Figure 6.**
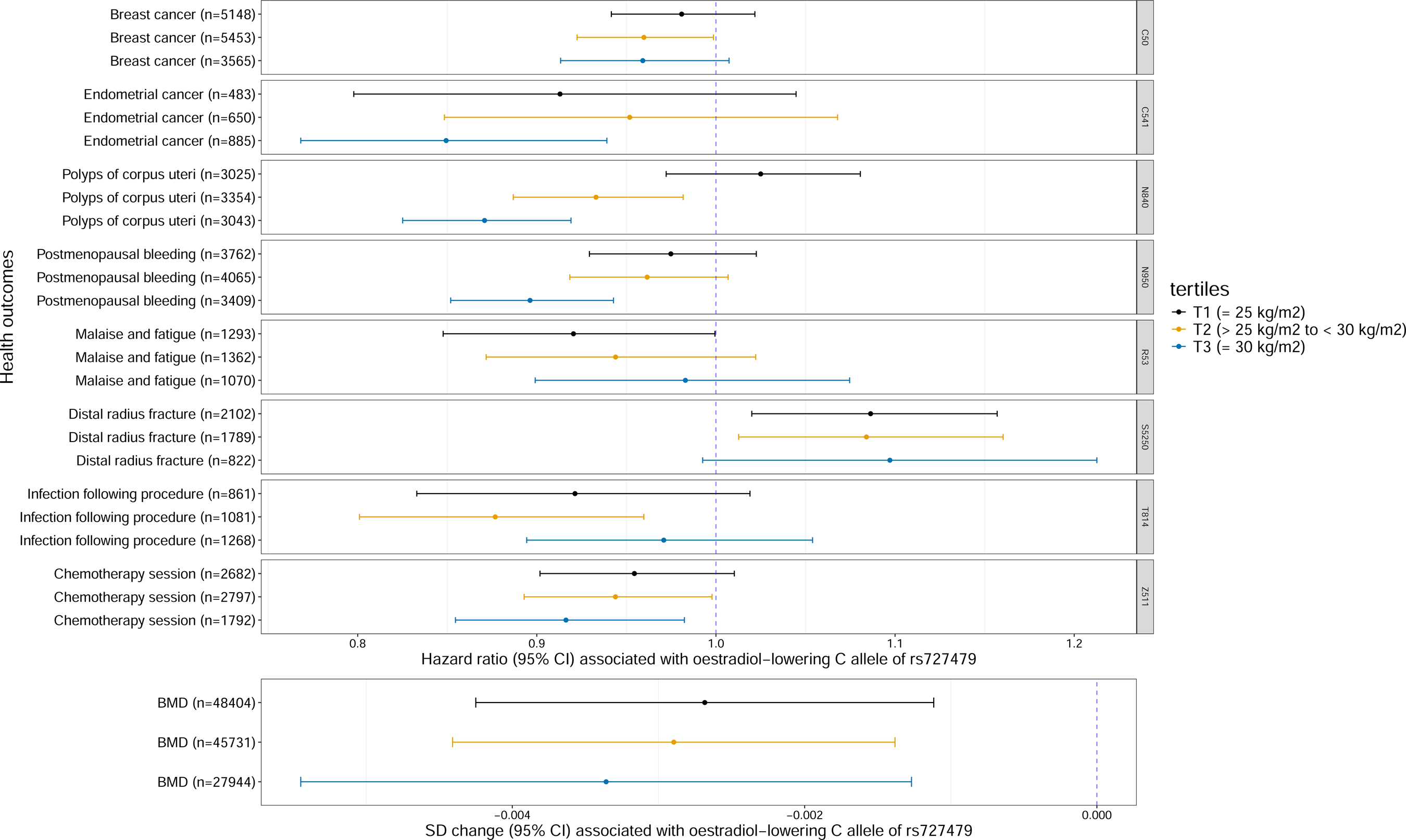
Association estimates for PheWAS outcomes and rs727479 across BMI tertiles. The estimates are in unit of hazard ratio (95% CI) for time-to-event outcomes or SD change (95% CI) for continuous outcome. The estimates have been scaled to reflect the effect of oestradiol-lowering C allele of rs727479 (mimicking aromatase inhibition). The y-axis has information on the health outcomes with the number of cases in each tertile (1st tertile (black): BMI ≤ 25 kg/m^2^, 2nd tertile (yellow): BMI ≥ 25 kg/m^2^ and < 30 kg/m^2^, 3rd tertile (blue): BMI ≥ 30 kg/m^2^)

## Discussion

In this study, we utilized rs727479, in the *CYP19A1* gene, to genetically proxy the long-term effects of aromatase inhibition. Our analyses recapitulate known effects of aromatase inhibition on reductions in ER+ breast cancer risk and bone mineral density but also identify novel long-term protective effects on endometrial cancer risk, supporting the repurposing potential of anastrozole to prevention of this cancer. While IBIS-II trial reported increased rates of hypertension, arthralgia, carpal tunnel syndrome, and influenza following five years of anastrozole treatment, we found no strong evidence for long-term effects of genetically-proxied aromatase inhibition on these outcomes. However, our estimates were imprecise and modest long-term effects cannot be excluded. In PheWAS analysis examining 449 disease and health-related outcomes not measured in IBIS-II, genetically-proxied aromatase inhibition was associated with lower risk of five outcomes including endometrial polyps and postmenopausal bleeding but an increased risk of distal radius fracture. Finally, in subgroup analyses of clinically relevant patient characteristics, we observed that genetically-proxied aromatase inhibition conferred a stronger protective effect against endometrial polyps and postmenopausal bleeding in women with higher BMI.

The observed protective effect on endometrial cancer risk is directionally concordant with IBIS-II, which reported fewer endometrial cancer cases in the anastrozole arm but with substantial uncertainty due to limited statistical power (5 vs 7 cases in anastrozole vs placebo arm, HR:0·72, 95% CI:0·18-2·65). Similarly, our findings are consistent with results from the ATAC trial which randomised 6,186 postmenopausal women with early-stage breast cancer and found lower rates of endometrial cancer in women in the anastrozole arm (OR:0·25, 95% CI:0·08-0·63), though numbers of cases were limited (6 vs 24 in anastrozole vs tamoxifen arm) and tamoxifen is known to increase endometrial cancer risk.^9,28^ Our findings are also consistent with prior mendelian randomisation (MR) and genetic epidemiological studies that reported a role of serum oestradiol and the *CYP19A1* locus in endometrial cancer development.^16,29,30^ Our subtype-stratified analyses suggested that the protective effect is stronger for endometrioid than non-endometrioid endometrial cancer. Collectively, these results highlight the potential for repurposing anastrozole for endometrial cancer prevention, which merits further evaluation in clinical trials.

The IBIS-II trial reported increased rates of hypertension, arthralgia, and carpal tunnel syndrome during the five-year active therapy period. However, the trial investigators noted that these side effects were more common in the first year of treatment, suggesting that they were unlikely to materially differ in the post-treatment period. In our analyses of genetically-proxied aromatase inhibition, we found weak evidence of an increased long-term risk of hypertension, directionally consistent with IBIS-II, but little evidence to support increased long-term risk of arthralgia and carpal tunnel syndrome. In PheWAS exploring possible novel long-term adverse effects of genetically-proxied aromatase inhibition, we identified an additional six disease and health-related outcomes associated with aromatase inhibition, five of which showed protective associations. These included protective associations with endometrial polyps and reductions in postmenopausal bleeding - both of which can be markers of subsequent endometrial cancer development - as well as a reduced risk of cancer chemotherapy sessions, likely reflecting reduced cancer incidence. We also found evidence of an increased risk of fracture, consistent with the accelerated bone mineral density loss experienced by postmenopausal women on aromatase inhibitors. While these analyses are exploratory and require replication, they highlight the utility of PheWAS for uncovering long-term consequences of drug-target perturbation beyond those captured in clinical trials.

In subgroup analyses, we found evidence for a more pronounced protective effect of genetically-proxied aromatase inhibition on endometrial polyps and postmenopausal bleeding in women with higher BMI. Given that excess adiposity is a major determinant of endometrial pathology and that aromatase expression in postmenopausal women is highest in adipose tissue, this pattern is biologically plausible.^31,32^ For other outcomes, there was little evidence of an interaction with BMI, testosterone-to-SHBG ratio, PRS for BMD and PRS for breast cancer. Although IBIS-II secondary analyses suggested greater risk benefit among women with higher baseline oestradiol-SHBG ratios, direct comparison was limited because oestradiol-SHBG ratios could not be reliably assessed for postmenopausal women in UK Biobank and because genetic estimates reflect lifelong exposure rather than effects of treatments initiated in midlife. Nonetheless, our findings do not discount the potential utility of hormonal biomarkers to identify women most likely to benefit from preventive aromatase inhibition.

Strengths of this study include the use of a genetic instrument within the *CYP19A1* gene that is strongly associated with oestradiol levels in postmenopausal women to mimic pharmacological aromatase inhibition. This approach enabled estimation of the long-term effects of aromatase inhibition while minimising confounding and bias. Where possible, findings were benchmarked against results from the IBIS-II trial, allowing validation of our genetic instrument. The use of summary genetic association data from several large GWAS consortia and individual-level participant data from the UK Biobank provided the statistical power to evaluate the cancer repurposing potential of anastrozole, identify secondary effects, and examine clinically relevant subgroups.

This study has several limitations. Genetic instruments proxy lifelong perturbation of a pharmacological target and may not fully replicate the pharmacokinetics and pharmacodynamics of aromatase inhibition over specific life-stages (e.g. post-menopausal period). Human genetics approaches can only estimate the on-target (i.e. target-mediated) effects of medications and cannot capture possible off-target, compound-specific effects of aromatase inhibitors. Although our instrument was biologically relevant and validated using IBIS-II outcomes, residual horizontal pleiotropy (i.e. the instrument influencing cancer risk through pathways independent to aromatase inhibition) cannot be fully ruled out. MR analyses of non-breast cancer outcomes relied on GWAS not restricted to postmenopausal women and, in some cases, included men, potentially limiting applicability of these findings to the target population (postmenopausal women). The IBIS-II trial was conducted in postmenopausal women at elevated baseline risk of breast cancer, whereas our analyses were not strictly restricted to this specific high-risk subgroup, which may further contribute to observed differences in effect estimates and magnitudes. Finally, analyses were performed primarily in individuals of European ancestry, which may limit generalisability to other populations.

Our analyses identified a protective effect of genetically-proxied aromatase inhibition on endometrial cancer risk that was of a 2·5-fold greater magnitude than the protective effect observed for breast cancer, the condition for which anastrozole is approved as preventive therapy. These findings are biologically plausible given the key role of oestradiol in endometrial carcinogenesis but require experimental validation using long-term follow-up data from clinical trials of anastrozole to confirm a preventive effect for endometrial cancer. Analysis of electronic health record (EHR) data could provide another opportunity to gain insight into the endometrial cancer preventive potential of anastrozole and to clarify patient subgroups most likely to benefit from this medication. In addition, evaluation of anastrozole use within EHR data could permit further assessment of whether anastrozole use confers a long-term increased risk of less serious adverse events reported in IBIS-II such as hypertension, or whether risk of these conditions is only increased during the active therapy period.

In conclusion, our genetic analyses confirm a protective role of genetically-proxied aromatase inhibition on breast cancer risk and suggest a pronounced protective effect of this medication on endometrial cancer risk. In the PheWAS analyses, we did not detect strong evidence for adverse effects beyond the established effect on bone mineral density, although limited power for some outcomes and the use of multiple testing correction mean that modest adverse effects cannot be ruled out. Subgroup analyses suggested that women with higher BMI may derive greater benefit from aromatase inhibition, warranting further investigation in clinical trial and electronic health record data to validate findings.

## Contributors

DR, RMM, PCH, JY, KSB and PS contributed to conceptualisation and design of this study. DR, TB and TOM contributed to the acquisition of data. The statistical analysis of the data was undertaken by DR and TB. The original draft was written by DR. All authors reviewed, contributed to, and approved the final version of the manuscript.

## Data Availability

UK Biobank data can be accessed through application made at http://ukbiobank.ac.uk/register-apply/. GWAS summary data are publicly available at IEU OpenGWAS (https://opengwas.io/) and the GWAS Catalog (https://www.ebi.ac.uk/gwas/).

## Acknowledgements

We thank the Endometrial Cancer Association Consortium (ECAC) for providing the summary-level data used in this study. Funding for the ECAC genome-wide association studies was provided by the National Health and Medical Research Council of Australia, the US National Institutes of Health, European Research Council, Wellcome Trust Centre for Human Genetics, and Cancer Research UK (see O’Mara et al. (2018)^22^, for details).

Funding: DR is supported by a Wellcome Trust PhD studentship (grant number 228303/Z/23/Z). RMM is a National Institute for Health Research Senior Investigator (NIHR202411). RMM and PCH are supported by a Cancer Research UK (C18281/A29019) programme grant (the Integrative Cancer Epidemiology Programme). RMM is also supported by the NIHR Bristol Biomedical Research Centre which is funded by the NIHR (BRC-1215-20011) and is a partnership between University Hospitals Bristol and Weston NHS Foundation Trust and the University of Bristol. RMM and PCH are affiliated with the Medical Research Council Integrative Epidemiology Unit at the University of Bristol which is supported by the Medical Research Council (MC_UU_00011/1, MC_UU_00011/3, MC_UU_00011/6, and MC_UU_00011/4) and the University of Bristol.

The funders had no role in study design, data collection, analysis, interpretation, manuscript writing, or the decision to submit the manuscript for publication.

## Data Sharing Statement

All analyses were undertaken using UK Biobank data (application number 81499) and publicly available GWAS summary statistics from IEU OpenGWAS (https://opengwas.io/) and the GWAS Catalog (https://www.ebi.ac.uk/gwas/). UK Biobank data are available through approved application. No new data was generated in this study.

