## Supplementary Methods for "Genetic prediction of long-term effects of aromatase inhibition on cancer and non-neoplastic disease risk"

This study was guided by a pre-specified research analysis plan submitted to the University of Bristol Pure repository prior to commencement of analyses.^1^ The analysis plan pre-specified the development and validation of a genetic instrument for aromatase inhibition, positive control analyses against established effects of aromatase inhibition, investigation of secondary disease outcomes, evaluation of cancer repurposing potential of aromatase inhibition, PheWAS, and subgroup analyses based on body mass index (BMI) and breast cancer PRS. Additional subgroup analyses, including those based on testosterone-to-SHBG ratio and PRS for BMD, were included after analyses had begun. These additional subgroup analyses were included following consultation with investigators from the IBIS-II trial to ensure clinical relevance and to extend evaluation to outcomes and patient characteristics not directly assessed in the original trial.^2^

**Mendelian randomisation analyses**

For genetic instrument validation and analyses evaluating the repurposing potential of aromatase inhibition and IBIS-reported adverse effects, the Mendelian randomisation (MR) approach was used. MR can generate unbiased estimates of causal effects if the genetic instrument: (i) associates with the exposure (relevance), (ii) is independent of confounders (exchangeability), and (iii) influences the outcome only through the exposure (exclusion restriction).^3^ Effect estimates and corresponding 95% confidence intervals (CIs) for genetically-proxied aromatase inhibition on outcomes of interest were calculated using the Wald ratio and delta method approximation and were scaled to reflect a one standard deviation (SD) decrease in serum oestradiol levels. The analyses were performed in R version 4.3.1 using the “TwoSampleMR” package.^4^ The IEU OpenGWAS^5^ and NHGRI-EBI GWAS Catalog^6^ databases were used to identify relevant GWAS to derive summary-level data for outcomes of interest.

**Identification of potential novel adverse effects of anastrozole**

To identify potential novel secondary effects of genetically-proxied aromatase inhibition, a phenome-wide association study (PheWAS) was conducted diseases and health-related outcomes (summary diagnoses that were clinically identified and self-reported illnesses) available in the UK Biobank. To perform the PheWAS, an open source tool called PHESANT (PHEnome Scan ANalysis Tool) was utilized.^7^ PHESANT is a software developed for performing automated tests of association of exposure of interest with a wide range of phenotypes in UK Biobank. Broadly, the implementation of PHESANT requires two input files, (a) phenotype file, and (b) a trait of interest file with information on the genetic variant(s).

To create the phenotype file, data corresponding to ICD-10 summary diagnoses (data-field 41270), self-reported non-cancer illness (data-field 20002) and self-reported cancer illness (data-field 20001) outcomes were extracted. Females were identified from the phenotype file using the “sex” data-field 31. This subset was then refined to include only postmenopausal women for the main PheWAS analysis, as anastrozole is primarily prescribed for high-risk postmenopausal women. Menopausal status was determined using data-field 2724 (“Had menopause”), yielding a final dataset of 162,360 women.

The UK Biobank genotype data was used to determine each individual’s dosage value for rs727479, ranging from 0 to 2, based on the expected number of copies of the effect allele they carried. This dosage information was integrated into the “trait of interest” file. Age at recruitment (data-field 21022) and the first 10 principal components (PCs) were identified as covariates for the analysis. To exclude less powered outcomes, only phenotypes with at least 1,000 cases were retained.

For comparability with our MR findings, the effect estimates from the PheWAS were converted to “MR” estimates by calculating a Wald ratio and corresponding standard error (SE) derived using the delta method.^8^ The p-values were adjusted for multiple testing using the Benjamini-Hochberg method^9^, with statistical “significance” defined as an FDR-adjusted p-value of less than 5%.

**Interactions with clinically relevant subgroups**

For each subgroup analysis, we first stratified participants into tertiles of the relevant clinical variable (i.e. BMI, testosterone-to-SHBG ratio, breast cancer PRS, BMD PRS). We then used Cox proportional hazards models to estimate hazard ratios for outcomes per SD increase in genetically-proxied aromatase inhibition in each tertile.^10^ Survival time for each participant was calculated in days from the year of birth to the year of diagnosis for the relevant health outcome, death, or end of follow-up period (corresponding to the latest data update for our UK Biobank project at the time of analysis: 07 June 2022), whichever came first. Welsh participants were followed up until diagnosis of health outcome, death or 31 December 2016, whichever came first, as cancer registry data for Welsh participants were only reliably linked up to this date and incident cases beyond this date might be incomplete. ^11^ In contrast to the time-to-event outcomes, heel BMD was evaluated using linear regression models fitted within each subgroup tertile to assess potential treatment heterogeneity. All models were adjusted for age at recruitment and the first 10 principal components of genetic ancestry.

To assess the statistical evidence for interaction, an interaction term between rs727479 and a continuous measure of the subgroup variable was included in the model and the fit of this model was compared to a null model (without the interaction term) using the log-likelihood ratio test (LRT). The *P*-values for the log-likelihood ratio estimates were adjusted for multiple testing using a Benjamini-Hochberg FDR correction.^9^

The biological relevance for each subgroup selection along with the details on stratification are outlined below.

**Body Mass Index (BMI)**

Aromatase activity in postmenopausal women is higher in adipose tissues as compared to other peripheral tissues, making adipose tissue a primary source of oestrogen production in this group.^12^ Consequently, women with greater body weight may have higher aromatase levels and thereby, may derive greater therapeutic benefits from aromatase inhibitors. However, elevated BMI may also increase the risk of aromatase inhibitor-related side effects. To test whether the effect of aromatase inhibition varied by BMI, a subgroup analysis was performed using data on BMI measures from UK Biobank (data field 21001). For comparability with the IBIS-II trial, participants were stratified into tertiles of BMI: ≤ 25 kg/m^2^, > 25 kg/m^2^ to < 30 kg/m^2^, and ≥ 30 kg/m^2^.

**Testosterone-to-sex-hormone binding globulin (SHBG) ratio**

Increased serum concentrations of sex hormones such as oestrogen and testosterone have been associated with an increased risk of breast cancer in postmenopausal women. ^13^ In contrast, higher levels of sex hormone-binding globulin (SHBG) have been associated with a reduced risk of breast cancer.^14^ However, the extent to which these hormones might influence the efficacy of aromatase inhibitors remains uncertain. A case-control study within the IBIS-II trial investigating the effects of serum oestradiol, testosterone, and SHBG levels on the efficacy of anastrozole in BC prevention observed that women with medium or high oestradiol-to-SHBG ratios had greater reduction in risk of BC with anastrozole as compared to women with low oestradiol-to-SHBG ratio.^15^ A similar but weaker association was observed for the testosterone-to-SHBG ratio.

To build on the findings of this case-control study and investigate the effects of aromatase inhibition across tertiles of this ratio, testosterone-to-SHBG ratio was identified as a subgroup of interest. We used testosterone-to-SHBG ratio as a surrogate for oestradiol-SHBG ratio because oestradiol concentrations in postmenopausal women in the UK Biobank are often below the detection limit of the immunoassay used.^16^ Testosterone and SHBG measurements were obtained from UK Biobank data-fields 30850 and 30830, respectively. For each participant, the ratio was calculated by dividing testosterone levels by SHBG concentrations and categorised into tertiles ensuring equal distribution in each tertile.

**Polygenic risk score (PRS) for bone mineral density (BMD)**

It is well established that reduction in oestrogen levels resulting from aromatase inhibition can lead to a loss in bone mineral density, given the key role of oestrogen in bone metabolism.^17,18^ As a result, individuals with greater genetic predisposition to low BMD may be at a higher risk of developing bone-related complications, such as fractures, following aromatase inhibition. . To assess if genetic predisposition influences the variability in outcomes associated with aromatase inhibition, we developed a polygenic risk score (PRS) for BMD using PRS-cs.^19^ A PRS was used in this case to avoid collider bias that could arise from stratifying on BMD, as rs727479 influences both aromatase activity and BMD. Specifically, the PRS was constructed for femoral neck BMD (FNBMD) using meta-analysed, females-only GWAS data from the Genetic Factors for Osteoporosis Consortium (GEFOS), a large genetic consortium osteoporotic study populations in Europe.^20^ The effect of genetically-proxied aromatase inhibition was then evaluated across tertiles of the BMD PRS.

**Breast cancer PRS**

Aromatase inhibitors target oestrogen synthesis, which plays an important role in hormone-sensitive breast cancer. To understand how an individual’s genetic risk of developing breast cancer might influence the therapeutic effect of aromatase inhibitors, PRS was developed using data from the Breast Cancer Association Consortium (BCAC) in PRS-cs.^21^ The PRS was then stratified into tertiles (ensuring equal number of individuals in each tertile) to assess the interaction between varying genetic predisposition to breast cancer and efficacy of genetically-proxied aromatase inhibition.
