## Supplementary Table 1 for "Genetic prediction of long-term effects of aromatase inhibition on cancer and non-neoplastic disease risk"

**Supplementary Table 1. MR estimates for IBIS-reported outcomes (including adverse events and cancer outcomes)**

| Outcome | Effect size (95% CI) |
| --- | --- |
| Genetic instrument validation | |
| ER+ breast cancer | 0.78 (0.67,0.92) |
| ER- breast cancer | 0.94 (0.74,1.20) |
| Overall breast cancer | 0.85 (0.74,0.97) |
| Heel BMD* | -0.32 (-0.36,-0.28) |
| Repurposing potential for other cancers | |
| Ovarian cancer | 0.84 (0.63,1.11) |
| Non-melanoma skin cancer | 0.83 (0.68,1.02) |
| Lung cancer | 1.15 (0.89,1.47) |
| Endometrial cancer | 0.30 (0.21,0.42) |
| Colorectal cancer | 0.98 (0.84,1.15) |
| ER- breast cancer | 0.94 (0.74,1.20) |
| IBIS-II reported adverse effects | |
| Influenza | 0.40 (0.22,0.73) |
| Carpal tunnel syndrome | 0.92 (0.67,1.27) |
| Arthralgia | 1.76 (0.82,3.78) |
| Hypertension | 1.09 (0.98,1.21) |

Effect sizes are reported as odds ratios per SD unit decrease in genetically-proxied serum oestradiol, with the exception of the continuous outcome* which is reported as 1-SD change per SD unit decrease in genetically-proxied serum oestradiol.
