## Supplementary Table 2 for "Genetic prediction of long-term effects of aromatase inhibition on cancer and non-neoplastic disease risk"

**Supplementary Table 2. PheWAS results for rs727479 and UK biobank health outcomes**

| Outcome | OR (95% CI) | P-value | Cases/Control |
| --- | --- | --- | --- |
| Polyp of corpus uteri | 0.58 (0.45,0.74) | 0.005 | 7550/154810 |
| Fracture of lower end of radius (closed) | 1.92 (1.39,2.66) | 0.020 | 3904/158456 |
| Chemotherapy session for neoplasm | 0.59 (0.45,0.77) | 0.023 | 5940/156420 |
| Postmenopausal bleeding | 0.67 (0.54,0.83) | 0.028 | 10011/152349 |
| Malaise and fatigue | 0.49 (0.33,0.72) | 0.028 | 2856/159504 |
| Infection following a procedure, not elsewhere classified | 0.46 (0.29,0.70) | 0.028 | 2327/160033 |
| Agranulocytosis | 0.53 (0.35,0.79) | 0.133 | 2617/159743 |
| Other antineoplastic drugs | 0.48 (0.30,0.77) | 0.133 | 1986/160374 |
| Hypothyroidism, unspecified | 0.76 (0.64,0.91) | 0.133 | 14973/147387 |
| Impingement syndrome of shoulder | 0.54 (0.35,0.82) | 0.164 | 2471/159889 |
| Rotator cuff syndrome | 0.52 (0.33,0.82) | 0.181 | 2170/160190 |
| Ascites | 0.44 (0.25,0.78) | 0.181 | 1348/161012 |
| Other specified noninflammatory disorders of uterus | 0.56 (0.38,0.84) | 0.181 | 2694/159666 |
| Falls (home) | 1.79 (1.18,2.70) | 0.191 | 2433/159927 |
| Septicaemia, unspecified | 0.62 (0.43,0.88) | 0.204 | 3489/158871 |
| Hiatus hernia* | 1.52 (1.12,2.06) | 0.204 | 4496/157844 |
| Intraductal carcinoma in situ | 0.47 (0.27,0.83) | 0.204 | 1422/160938 |
| Pneumonia, unspecified | 0.62 (0.43,0.89) | 0.204 | 3414/158946 |
| Other iron deficiency anaemias | 0.55 (0.35,0.87) | 0.204 | 2101/160259 |
| Other specified disorders of bone density and structure | 1.77 (1.14,2.76) | 0.204 | 2130/160230 |
| Benign neoplasm of breast | 0.43 (0.22,0.82) | 0.204 | 1012/161348 |
| Diaphragmatic hernia without obstruction or gangrene | 1.22 (1.05,1.43) | 0.204 | 19509/142851 |
| Living alone | 0.55 (0.35,0.88) | 0.204 | 2056/160304 |
| Chronic renal failure, unspecified | 0.61 (0.41,0.90) | 0.204 | 2889/159471 |
| Follow-up care involving removal of fracture plate and other internal fixation device | 1.87 (1.15,3.04) | 0.204 | 1737/160623 |
| Unspecified haemorrhoids with other complications | 2.09 (1.17,3.71) | 0.204 | 1240/161120 |
| Chronic obstructive pulmonary disease with acute lower respiratory infection | 1.75 (1.13,2.72) | 0.204 | 2161/160199 |
| Personal history of psychoactive substance abuse | 0.83 (0.71,0.97) | 0.274 | 20660/141700 |
| Arthritis, unspecified (Site unspecified) | 0.63 (0.42,0.93) | 0.307 | 2755/159605 |
| Other specified soft tissue disorders (Lower leg) | 0.61 (0.40,0.93) | 0.307 | 2382/159978 |
| Spine arthritis/spondylitis* | 1.78 (1.09,2.90) | 0.307 | 1714/160626 |
| Uterine fibroids* | 1.55 (1.07,2.25) | 0.307 | 2978/159362 |
| Personal history of malignant neoplasm of genital organs | 0.60 (0.38,0.93) | 0.315 | 2168/160192 |
| Peritoneal adhesions | 0.63 (0.42,0.94) | 0.325 | 2648/159712 |
| Haemorrhoids, unspecified | 0.66 (0.46,0.95) | 0.326 | 3307/159053 |
| Personal history of malignant neoplasm of breast | 0.79 (0.63,0.98) | 0.393 | 9492/152868 |
| Disorientation, unspecified | 0.62 (0.40,0.96) | 0.393 | 2302/160058 |
| Low back pain (Site unspecified) | 0.50 (0.26,0.95) | 0.406 | 1042/161318 |
| Axillary and upper limb lymph nodes | 0.64 (0.43,0.97) | 0.406 | 2569/159791 |
| Myopia | 0.57 (0.34,0.96) | 0.406 | 1579/160781 |
| Hypokalaemia | 0.65 (0.44,0.98) | 0.408 | 2730/159630 |
| Atrial fibrillation and flutter | 0.74 (0.55,0.99) | 0.434 | 5330/157030 |
| Secondary malignant neoplasm of liver | 0.66 (0.44,0.99) | 0.452 | 2571/159789 |
| Localised enlarged lymph nodes | 0.55 (0.31,0.99) | 0.452 | 1271/161089 |
| Presence of orthopaedic joint implants | 1.23 (1.00,1.50) | 0.452 | 11431/150929 |
| Benign neoplasm of ovary | 0.57 (0.33,0.99) | 0.465 | 1413/160947 |
| Other specified intervertebral disk degeneration | 0.63 (0.40,1.00) | 0.465 | 2092/160268 |
| Deficiency of other specified B group vitamins | 0.53 (0.28,1.00) | 0.478 | 1061/161299 |
| Unstable angina | 0.61 (0.37,1.01) | 0.480 | 1755/160605 |
| Coeliac disease | 0.58 (0.33,1.01) | 0.480 | 1347/161013 |
| Osteoporosis, unspecified | 1.25 (0.99,1.57) | 0.480 | 8483/153877 |
| Emphysema, unspecified | 1.69 (0.99,2.89) | 0.480 | 1430/160930 |
| Personal history of diseases of the musculoskeletal system and connective tissue | 0.63 (0.39,1.01) | 0.480 | 1871/160489 |
| Cholelithiasis/gall stones* | 1.37 (0.99,1.89) | 0.482 | 4057/158283 |
| Other surgical procedures | 0.68 (0.45,1.02) | 0.490 | 2613/159747 |
| Arthritis, unspecified (Multiple sites) | 0.55 (0.30,1.03) | 0.490 | 1109/161251 |
| Angina pectoris, unspecified | 1.27 (0.98,1.64) | 0.520 | 6666/155694 |
| Fall (Unspecified place) | 1.59 (0.97,2.60) | 0.520 | 1690/160670 |
| Calculus of kidney | 1.69 (0.95,3.00) | 0.557 | 1241/161119 |
| Personal history of irradiation | 0.74 (0.54,1.03) | 0.557 | 4052/158308 |
| Appendicitis* | 0.61 (0.35,1.06) | 0.557 | 1427/160913 |
| Secondary malignant neoplasm of bone and bone marrow | 0.65 (0.40,1.05) | 0.557 | 1880/160480 |
| Gestational hypertension/pre-eclampsia* | 0.57 (0.30,1.07) | 0.557 | 1062/161278 |
| Transient cerebral ischaemic attack, unspecified | 1.67 (0.94,2.97) | 0.557 | 1249/161111 |
| Chronic kidney disease, stage 3 | 0.75 (0.55,1.04) | 0.557 | 4370/157990 |
| Personal history of chemotherapy for neoplastic disease | 0.76 (0.56,1.04) | 0.570 | 4444/157916 |
| Delirium, unspecified | 0.62 (0.36,1.07) | 0.570 | 1438/160922 |
| Endometriosis* | 1.58 (0.92,2.71) | 0.633 | 1429/160911 |
| Unknown and unspecified causes of morbidity | 1.20 (0.97,1.48) | 0.653 | 9782/152578 |
| Acquired absence of breast(s) | 0.75 (0.53,1.06) | 0.663 | 3577/158783 |
| Dental caries, unspecified | 1.55 (0.91,2.62) | 0.663 | 1490/160870 |
| Glaucoma, unspecified | 0.73 (0.50,1.07) | 0.673 | 3049/159311 |
| Other chest pain | 1.27 (0.95,1.70) | 0.673 | 4994/157366 |
| Coxarthrosis, unspecified | 1.20 (0.96,1.51) | 0.673 | 8523/153837 |
| Glaucoma suspect | 0.61 (0.34,1.12) | 0.673 | 1181/161179 |
| Aortic (valve) stenosis | 1.54 (0.90,2.64) | 0.673 | 1430/160930 |
| Unspecified fall (Unspecified place) | 1.38 (0.92,2.06) | 0.673 | 2608/159752 |
| Trigger finger | 0.63 (0.35,1.13) | 0.673 | 1263/161097 |
| Hypo-osmolality and hyponatraemia | 0.75 (0.52,1.08) | 0.673 | 3359/159001 |
| Special screening examination, unspecified | 1.43 (0.91,2.24) | 0.683 | 2078/160282 |
| Other specified disorders of eyelid | 0.64 (0.36,1.13) | 0.686 | 1318/161042 |
| Gonarthrosis, unspecified | 1.17 (0.96,1.42) | 0.691 | 11658/150702 |
| Other specified diseases of stomach and duodenum | 1.46 (0.90,2.36) | 0.696 | 1772/160588 |
| Secondary malignant neoplasm of retroperitoneum and peritoneum | 0.68 (0.41,1.12) | 0.696 | 1678/160682 |
| Personal history of long-term (current) use of other medicaments | 1.16 (0.96,1.41) | 0.696 | 11987/150373 |
| Rheumatoid arthritis, unspecified (Site unspecified) | 0.68 (0.40,1.13) | 0.707 | 1618/160742 |
| Hearing loss, unspecified | 0.76 (0.53,1.10) | 0.707 | 3179/159181 |
| Dermatitis, unspecified | 0.66 (0.38,1.15) | 0.707 | 1382/160978 |
| Hallux valgus (acquired) | 1.22 (0.93,1.59) | 0.707 | 6068/156292 |
| Psoriasis, unspecified | 0.64 (0.35,1.16) | 0.707 | 1187/161173 |
| Personal history of allergy to penicillin | 0.87 (0.72,1.05) | 0.707 | 12934/149426 |
| Osteoarthritis* | 1.13 (0.96,1.32) | 0.707 | 18955/143385 |
| Pneumonia* | 0.73 (0.47,1.13) | 0.720 | 2224/160116 |
| Fibromyalgia | 1.52 (0.86,2.68) | 0.720 | 1274/161086 |
| Special screening examination for neoplasm of intestinal tract | 1.28 (0.91,1.80) | 0.720 | 3700/158660 |
| Senile incipient cataract | 0.72 (0.46,1.13) | 0.720 | 2133/160227 |
| Polyp of colon | 0.84 (0.67,1.07) | 0.724 | 8082/154278 |
| Cardiac murmur, unspecified | 1.51 (0.85,2.67) | 0.724 | 1267/161093 |
| Arthrosis, unspecified (Site unspecified) | 0.78 (0.56,1.10) | 0.734 | 3661/158699 |
| Diabetes* | 0.80 (0.59,1.09) | 0.734 | 4564/157776 |
| Acute myocardial infarction, unspecified | 1.56 (0.83,2.91) | 0.739 | 1057/161303 |
| Other primary gonarthrosis | 1.33 (0.89,1.99) | 0.742 | 2580/159780 |
| Gastro-oesophageal reflux disease with oesophagitis | 1.20 (0.92,1.56) | 0.742 | 6285/156075 |
| Special screening examination for diseases of the blood and blood-forming organs and certain disorders involving the immune mechanism | 0.72 (0.45,1.15) | 0.753 | 1966/160394 |
| Lesion of plantar nerve | 0.65 (0.35,1.22) | 0.757 | 1095/161265 |
| Ganglion | 0.71 (0.43,1.17) | 0.757 | 1738/160622 |
| Other disorders of lung | 0.66 (0.36,1.21) | 0.757 | 1173/161187 |
| Left bundle-branch block, unspecified | 0.70 (0.42,1.18) | 0.757 | 1599/160761 |
| Other complications of procedures, not elsewhere classified | 0.67 (0.37,1.21) | 0.757 | 1219/161141 |
| Raynaud's syndrome | 1.44 (0.83,2.50) | 0.768 | 1380/160980 |
| Ptosis of eyelid | 1.51 (0.81,2.81) | 0.768 | 1078/161282 |
| Lobar pneumonia, unspecified | 1.21 (0.91,1.61) | 0.768 | 5200/157160 |
| Seborrhoeic keratosis | 1.33 (0.86,2.06) | 0.768 | 2230/160130 |
| Urinary tract infection, site not specified | 1.15 (0.93,1.42) | 0.769 | 9983/152377 |
| Disorder of skin and subcutaneous tissue, unspecified | 1.28 (0.88,1.85) | 0.769 | 3059/159301 |
| Other ill-defined heart diseases | 1.43 (0.83,2.46) | 0.769 | 1396/160964 |
| Allergic rhinitis due to pollen | 0.67 (0.37,1.24) | 0.769 | 1152/161208 |
| Other and unspecified intestinal obstruction | 0.72 (0.43,1.20) | 0.769 | 1608/160752 |
| Osteoporosis* | 1.20 (0.91,1.58) | 0.769 | 5570/156770 |
| Umbilical hernia without obstruction or gangrene | 1.50 (0.80,2.79) | 0.769 | 1063/161297 |
| Deep venous thrombosis (dvt)* | 0.81 (0.58,1.13) | 0.790 | 3764/158576 |
| Ulcerative colitis, unspecified | 0.70 (0.40,1.23) | 0.790 | 1342/161018 |
| Paroxysmal atrial fibrillation | 0.76 (0.49,1.18) | 0.790 | 2204/160156 |
| Other specified urinary incontinence | 1.43 (0.81,2.52) | 0.790 | 1286/161074 |
| Staphylococcus aureus as the cause of diseases classified to other chapters | 0.69 (0.38,1.25) | 0.793 | 1179/161181 |
| Street and highway | 0.72 (0.42,1.22) | 0.804 | 1525/160835 |
| Osteoporosis, unspecified (Site unspecified) | 1.23 (0.88,1.71) | 0.811 | 3852/158508 |
| Senility | 0.69 (0.38,1.26) | 0.811 | 1202/161158 |
| Other specified diseases of anus and rectum | 0.73 (0.43,1.23) | 0.814 | 1572/160788 |
| Intestinal bypass and anastomosis status | 0.73 (0.44,1.23) | 0.814 | 1598/160762 |
| Eczema/dermatitis* | 0.82 (0.58,1.15) | 0.814 | 3793/158547 |
| Abnormal findings on diagnostic imaging of other parts of digestive tract | 1.41 (0.79,2.54) | 0.814 | 1215/161145 |
| Fall on and from stairs and steps (Home) | 0.75 (0.46,1.22) | 0.814 | 1751/160609 |
| Cellulitis of other parts of limb | 0.80 (0.55,1.17) | 0.814 | 3059/159301 |
| Escherichia coli [E. Coli] as the cause of diseases classified to other chapters | 1.24 (0.86,1.80) | 0.814 | 3085/159275 |
| Polymyalgia rheumatica | 1.32 (0.82,2.12) | 0.814 | 1867/160493 |
| Unspecified urinary incontinence | 1.27 (0.85,1.91) | 0.814 | 2520/159840 |
| Palliative care | 0.83 (0.60,1.14) | 0.824 | 4210/158150 |
| Pure hypercholesterolaemia | 0.91 (0.78,1.07) | 0.828 | 20014/142346 |
| Epistaxis | 1.37 (0.79,2.37) | 0.833 | 1387/160973 |
| Fever, unspecified | 0.80 (0.54,1.19) | 0.838 | 2710/159650 |
| Stroke* | 1.32 (0.81,2.15) | 0.858 | 1740/160600 |
| Atherosclerotic heart disease | 1.15 (0.89,1.49) | 0.878 | 6632/155728 |
| Fractures of other parts of lower leg (closed) | 1.32 (0.79,2.21) | 0.878 | 1592/160768 |
| Peripheral vascular disease, unspecified | 1.36 (0.78,2.36) | 0.879 | 1349/161011 |
| Depressive episode, unspecified | 0.89 (0.72,1.10) | 0.879 | 10274/152086 |
| Female pelvic peritoneal adhesions | 1.32 (0.79,2.19) | 0.879 | 1610/160750 |
| Bronchitis* | 0.73 (0.41,1.30) | 0.879 | 1293/161047 |
| Arthrosis, unspecified (Shoulder region) | 0.76 (0.45,1.27) | 0.879 | 1557/160803 |
| Parkinson's disease | 0.71 (0.37,1.35) | 0.879 | 1007/161353 |
| Phlebitis and thrombophlebitis of other deep vessels of lower extremities | 0.77 (0.47,1.27) | 0.890 | 1693/160667 |
| Other spondylosis (Lumbar region) | 1.30 (0.78,2.16) | 0.890 | 1626/160734 |
| Pulmonary embolism without mention of acute cor pulmonale | 1.22 (0.83,1.81) | 0.890 | 2763/159597 |
| Irritable bowel syndrome* | 0.86 (0.64,1.15) | 0.890 | 4961/157379 |
| Thrombocytopenia, unspecified | 1.39 (0.73,2.65) | 0.890 | 1004/161356 |
| High cholesterol* | 0.92 (0.78,1.08) | 0.890 | 18898/143442 |
| Spondylosis, unspecified | 0.75 (0.43,1.31) | 0.890 | 1380/160980 |
| Insulin-dependent diabetes mellitus (Without complications) | 0.72 (0.39,1.36) | 0.890 | 1066/161294 |
| Family history of malignant neoplasm of digestive organs | 1.16 (0.87,1.55) | 0.890 | 5020/157340 |
| Other gastritis | 1.16 (0.86,1.57) | 0.890 | 4768/157592 |
| Other fall on same level (Home) | 0.74 (0.40,1.36) | 0.890 | 1160/161200 |
| Follow-up care involving plastic surgery of breast | 0.78 (0.47,1.29) | 0.890 | 1697/160663 |
| Other and unspecified symptoms and signs involving the urinary system | 0.74 (0.41,1.37) | 0.890 | 1155/161205 |
| Cerebral infarction, unspecified | 1.29 (0.77,2.16) | 0.890 | 1561/160799 |
| Dyspnoea | 0.87 (0.65,1.16) | 0.890 | 5034/157326 |
| Chest pain, unspecified | 1.10 (0.90,1.34) | 0.890 | 11353/151007 |
| Transverse colon | 0.79 (0.48,1.29) | 0.890 | 1782/160578 |
| Allergy/hypersensitivity/anaphylaxis* | 1.30 (0.75,2.26) | 0.890 | 1372/160968 |
| Heart attack/myocardial infarction* | 0.78 (0.46,1.31) | 0.890 | 1592/160748 |
| Unspecified haematuria | 1.13 (0.87,1.47) | 0.890 | 6347/156013 |
| Fracture of neck of femur (closed) | 0.80 (0.50,1.27) | 0.890 | 1992/160368 |
| Melaena | 1.30 (0.75,2.24) | 0.890 | 1400/160960 |
| Dyspepsia | 0.89 (0.69,1.14) | 0.890 | 7133/155227 |
| Personal history of allergy to other drugs, medicaments and biological substances | 0.89 (0.69,1.14) | 0.890 | 7217/155143 |
| Stress incontinence | 0.86 (0.62,1.18) | 0.890 | 4211/158149 |
| Trichilemmal cyst | 1.23 (0.79,1.90) | 0.890 | 2205/160155 |
| Heart valve problem/heart murmur* | 1.30 (0.74,2.31) | 0.890 | 1271/161069 |
| Cervical spondylosis* | 1.28 (0.75,2.17) | 0.890 | 1488/160852 |
| Back problem* | 0.83 (0.56,1.24) | 0.890 | 2681/159659 |
| Irritable bowel syndrome without diarrhoea | 0.88 (0.66,1.17) | 0.890 | 5199/157161 |
| Volume depletion | 0.85 (0.60,1.21) | 0.890 | 3421/158939 |
| Muscle/soft tissue problem* | 0.78 (0.45,1.36) | 0.890 | 1392/160948 |
| Chronic obstructive pulmonary disease, unspecified | 1.13 (0.86,1.47) | 0.890 | 6061/156299 |
| Personal history of other specified conditions | 0.75 (0.39,1.43) | 0.890 | 1018/161342 |
| Internal haemorrhoids without complication | 0.80 (0.49,1.31) | 0.890 | 1774/160586 |
| Procedure not carried out because of patient's decision for other and unspecified reasons | 0.76 (0.41,1.40) | 0.890 | 1147/161213 |
| Rheumatoid arthritis, unspecified | 0.84 (0.56,1.24) | 0.890 | 2739/159621 |
| Ovarian cyst or cysts* | 0.82 (0.53,1.27) | 0.890 | 2255/160085 |
| Radiotherapy session | 0.77 (0.43,1.39) | 0.890 | 1225/161135 |
| Asthma* | 0.93 (0.79,1.09) | 0.890 | 18619/143721 |
| Varicose veins of lower extremities without ulcer or inflammation | 0.89 (0.69,1.16) | 0.890 | 6356/156004 |
| Psoriasis* | 0.80 (0.48,1.33) | 0.890 | 1597/160743 |
| Other chemotherapy | 1.13 (0.86,1.49) | 0.890 | 5533/156827 |
| Unclassifiable* | 0.90 (0.70,1.15) | 0.890 | 6859/155481 |
| Dizziness and giddiness | 1.15 (0.83,1.60) | 0.890 | 4003/158357 |
| Chronic ischaemic heart disease, unspecified | 1.12 (0.86,1.48) | 0.890 | 5914/156446 |
| Palpitations | 1.17 (0.81,1.67) | 0.890 | 3260/159100 |
| Rectal polyp | 0.87 (0.62,1.22) | 0.890 | 3741/158619 |
| Other obesity | 0.80 (0.46,1.37) | 0.890 | 1436/160924 |
| Basal cell carcinoma* | 0.82 (0.51,1.32) | 0.890 | 1894/160446 |
| Abnormal uterine and vaginal bleeding, unspecified | 1.30 (0.70,2.41) | 0.890 | 1086/161274 |
| Anxiety disorder, unspecified | 0.90 (0.70,1.15) | 0.890 | 7163/155197 |
| Excessive and frequent menstruation with regular cycle | 1.17 (0.81,1.68) | 0.890 | 3194/159166 |
| Other specified intervertebral disk displacement | 1.24 (0.74,2.05) | 0.890 | 1629/160731 |
| Oesophageal obstruction | 0.78 (0.43,1.41) | 0.890 | 1210/161150 |
| Other and unspecified convulsions | 0.78 (0.43,1.42) | 0.890 | 1186/161174 |
| Fitting and adjustment of urinary device | 0.76 (0.40,1.47) | 0.890 | 1002/161358 |
| Senile cataract, unspecified | 0.80 (0.47,1.37) | 0.890 | 1475/160885 |
| Obesity, unspecified | 1.09 (0.88,1.35) | 0.890 | 9903/152457 |
| Diverticular disease/diverticulitis* | 1.18 (0.78,1.80) | 0.890 | 2421/159919 |
| Procedure not carried out for other reasons | 1.09 (0.87,1.37) | 0.890 | 8702/153658 |
| Family history of malignant neoplasm of breast | 0.82 (0.49,1.36) | 0.890 | 1643/160717 |
| Other examinations for administrative purposes | 0.81 (0.47,1.39) | 0.890 | 1461/160899 |
| Syncope and collapse | 1.11 (0.85,1.46) | 0.890 | 5822/156538 |
| Personal history of malignant neoplasm of digestive organs | 0.87 (0.61,1.24) | 0.890 | 3376/158984 |
| Acquired absence of genital organ(s) | 0.90 (0.70,1.17) | 0.890 | 6516/155844 |
| Epidermal cyst | 1.27 (0.69,2.33) | 0.890 | 1113/161247 |
| Other spondylosis (Cervical region) | 0.84 (0.53,1.33) | 0.890 | 2035/160325 |
| Personal history of malignant neoplasm of urinary tract | 0.79 (0.43,1.45) | 0.890 | 1170/161190 |
| Fatty (change of) liver, not elsewhere classified | 1.20 (0.74,1.95) | 0.890 | 1778/160582 |
| Migraine, unspecified | 0.86 (0.57,1.28) | 0.890 | 2633/159727 |
| Astigmatism | 0.85 (0.55,1.32) | 0.890 | 2199/160161 |
| Family history of eye and ear disorders | 1.20 (0.73,1.95) | 0.890 | 1750/160610 |
| Other specified medical care | 0.79 (0.41,1.51) | 0.890 | 1001/161359 |
| Urethral stricture, unspecified | 0.81 (0.46,1.43) | 0.890 | 1313/161047 |
| Gastritis, unspecified | 0.93 (0.75,1.14) | 0.890 | 10270/152090 |
| Follow-up examination after surgery for malignant neoplasm | 1.18 (0.75,1.87) | 0.890 | 1998/160362 |
| Emphysema/chronic bronchitis* | 1.17 (0.76,1.81) | 0.890 | 2215/160125 |
| Sciatica* | 1.21 (0.72,2.04) | 0.890 | 1528/160812 |
| Anaemia, unspecified | 0.92 (0.74,1.15) | 0.890 | 9356/153004 |
| Arthritis (nos)* | 0.83 (0.50,1.39) | 0.890 | 1605/160735 |
| Other fecal abnormalities | 0.85 (0.53,1.35) | 0.890 | 1976/160384 |
| Other and unspecified symptoms and signs involving the nervous and musculoskeletal systems | 1.22 (0.70,2.15) | 0.890 | 1323/161037 |
| Abnormal findings on diagnostic imaging of lung | 1.17 (0.75,1.82) | 0.890 | 2188/160172 |
| Other and unspecified abnormalities of gait and mobility | 0.88 (0.61,1.27) | 0.890 | 3152/159208 |
| Senile nuclear cataract | 1.09 (0.86,1.37) | 0.890 | 8348/154012 |
| Alzheimer's disease, unspecified | 0.80 (0.42,1.51) | 0.890 | 1040/161320 |
| Special screening examination for other viral diseases | 1.09 (0.85,1.41) | 0.890 | 6821/155539 |
| Abnormal results of liver function studies | 0.88 (0.60,1.28) | 0.890 | 3036/159324 |
| Hayfever/allergic rhinitis* | 0.92 (0.73,1.16) | 0.890 | 8181/154159 |
| Nausea and vomiting | 0.93 (0.75,1.15) | 0.890 | 9795/152565 |
| Acidosis | 0.81 (0.43,1.51) | 0.890 | 1092/161268 |
| Superficial injury of other parts of head | 1.24 (0.66,2.32) | 0.890 | 1053/161307 |
| Diverticular disease of intestine, part unspecified, without perforation or abscess | 1.09 (0.84,1.42) | 0.890 | 6460/155900 |
| Other hammer toe(s) (acquired) | 1.17 (0.73,1.87) | 0.890 | 1933/160427 |
| Other and unspecified abdominal pain | 0.93 (0.76,1.15) | 0.890 | 10139/152221 |
| Low back pain | 1.14 (0.77,1.71) | 0.890 | 2633/159727 |
| Mixed anxiety and depressive disorder | 0.84 (0.49,1.42) | 0.890 | 1505/160855 |
| Uterine polyps* | 1.23 (0.66,2.31) | 0.890 | 1060/161280 |
| Personal history of allergy, other than to drugs and biological substances | 0.91 (0.67,1.22) | 0.890 | 4886/157474 |
| Personal history of malignant neoplasms of other organs and systems | 1.10 (0.83,1.47) | 0.890 | 5230/157130 |
| Personal history of diseases of the respiratory system | 0.85 (0.51,1.40) | 0.890 | 1675/160685 |
| Constipation | 0.93 (0.76,1.15) | 0.890 | 10238/152122 |
| Pain in limb (Lower leg) | 0.86 (0.55,1.35) | 0.890 | 2089/160271 |
| Other deformities of toe(s) (acquired) | 0.85 (0.51,1.41) | 0.890 | 1641/160719 |
| Rectum | 1.16 (0.73,1.84) | 0.890 | 1996/160364 |
| Nosocomial condition | 1.18 (0.71,1.96) | 0.890 | 1622/160738 |
| Calculus of gallbladder without cholecystitis | 0.92 (0.71,1.19) | 0.890 | 6460/155900 |
| Acute subendocardial myocardial infarction | 0.82 (0.45,1.51) | 0.890 | 1138/161222 |
| Personal history of allergy to other antibiotic agents | 0.91 (0.67,1.22) | 0.890 | 4887/157473 |
| Rash and other nonspecific skin eruption | 0.84 (0.48,1.45) | 0.890 | 1394/160966 |
| Arthrosis, unspecified (Hand) | 1.19 (0.70,2.02) | 0.890 | 1479/160881 |
| Other and unspecified lesions of oral mucosa | 0.83 (0.46,1.50) | 0.890 | 1214/161146 |
| Orthostatic hypotension | 1.17 (0.71,1.92) | 0.890 | 1716/160644 |
| Irregular menstruation, unspecified | 0.82 (0.44,1.53) | 0.890 | 1082/161278 |
| Asthma, unspecified | 0.95 (0.80,1.12) | 0.890 | 15652/146708 |
| Sleep apnoea | 1.17 (0.71,1.95) | 0.890 | 1629/160731 |
| Haemoptysis | 1.22 (0.64,2.33) | 0.890 | 1003/161357 |
| Removal of other organ (partial) (total) | 1.12 (0.77,1.64) | 0.890 | 2993/159367 |
| Tobacco use | 0.90 (0.63,1.27) | 0.890 | 3505/158855 |
| Accidental puncture and laceration during a procedure, not elsewhere classified | 0.83 (0.45,1.53) | 0.890 | 1122/161238 |
| Procedure not carried out because of contraindication | 0.91 (0.68,1.22) | 0.890 | 5112/157248 |
| Angina | 1.11 (0.78,1.58) | 0.890 | 3522/158818 |
| Depression* | 0.94 (0.76,1.16) | 0.890 | 10362/151978 |
| Headache | 1.09 (0.82,1.46) | 0.890 | 5072/157288 |
| Hemiplegia, unspecified | 1.19 (0.66,2.17) | 0.890 | 1162/161198 |
| Observation for other suspected diseases and conditions | 1.10 (0.80,1.50) | 0.890 | 4373/157987 |
| Epilepsy* | 0.84 (0.46,1.52) | 0.890 | 1199/161141 |
| Supraventricular tachycardia | 0.86 (0.52,1.43) | 0.890 | 1641/160719 |
| Viral infection, unspecified | 0.83 (0.44,1.57) | 0.890 | 1042/161318 |
| Follow-up examination after surgery for other conditions | 0.89 (0.60,1.32) | 0.890 | 2790/159570 |
| Polyp of stomach and duodenum | 1.09 (0.81,1.47) | 0.890 | 4778/157582 |
| Fracture of upper end of humerus (closed) | 1.20 (0.64,2.24) | 0.890 | 1063/161297 |
| Gastroenteritis and colitis of unspecified origin | 1.08 (0.83,1.39) | 0.890 | 6709/155651 |
| Abnormal weight loss | 1.09 (0.81,1.45) | 0.890 | 5178/157182 |
| Single live birth | 0.86 (0.49,1.49) | 0.890 | 1710/160650 |
| Pulmonary collapse | 1.12 (0.75,1.67) | 0.890 | 2652/159708 |
| Personal history of long-term (current) use of anticoagulants | 0.94 (0.75,1.17) | 0.890 | 9107/153253 |
| Scar conditions and fibrosis of skin | 0.86 (0.50,1.48) | 0.890 | 1451/160909 |
| Exposure to unspecified factor (Unspecified place) | 0.88 (0.54,1.42) | 0.890 | 1844/160516 |
| Non-insulin-dependent diabetes mellitus (Without complications) | 1.06 (0.85,1.32) | 0.890 | 9323/153037 |
| Special screening examination for other specified diseases and disorders | 0.86 (0.50,1.49) | 0.890 | 1428/160932 |
| Deviated nasal septum | 1.18 (0.64,2.17) | 0.890 | 1122/161238 |
| Low back pain (Lumbar region) | 1.16 (0.67,1.99) | 0.890 | 1422/160938 |
| Mitral (valve) insufficiency | 1.13 (0.71,1.81) | 0.890 | 1943/160417 |
| After-cataract | 1.13 (0.72,1.79) | 0.890 | 2018/160342 |
| Other specified disorders of nose and nasal sinuses | 1.18 (0.63,2.21) | 0.890 | 1068/161292 |
| Pleural effusion, not elsewhere classified | 1.08 (0.80,1.46) | 0.890 | 4771/157589 |
| Gastro-oesophageal reflux disease without oesophagitis | 0.95 (0.80,1.14) | 0.890 | 14578/147782 |
| Vitamin D deficiency, unspecified | 1.13 (0.71,1.81) | 0.890 | 1902/160458 |
| Ulcer of oesophagus | 1.13 (0.72,1.76) | 0.890 | 2126/160234 |
| Presence of coronary angioplasty implant and graft | 1.12 (0.72,1.74) | 0.890 | 2185/160175 |
| Nonspecific low blood-pressure reading | 1.18 (0.63,2.18) | 0.890 | 1092/161268 |
| Flatulence and related conditions | 1.15 (0.67,1.96) | 0.890 | 1471/160889 |
| Other and unspecified ovarian cysts | 0.91 (0.62,1.33) | 0.890 | 2920/159440 |
| Bladder problem (not cancer)* | 1.16 (0.65,2.06) | 0.890 | 1272/161068 |
| Secondary malignant neoplasm of lung | 0.88 (0.55,1.44) | 0.890 | 1816/160544 |
| Follow-up examination after unspecified treatment for other conditions | 1.15 (0.66,1.99) | 0.890 | 1395/160965 |
| Cardiomegaly | 0.90 (0.59,1.37) | 0.890 | 2459/159901 |
| Other complications of internal orthopaedic prosthetic devices, implants and grafts | 0.87 (0.49,1.54) | 0.890 | 1265/161095 |
| Observation for suspected myocardial infarction | 1.17 (0.62,2.22) | 0.890 | 1026/161334 |
| Presence of cardiac pacemaker | 0.88 (0.51,1.50) | 0.890 | 1477/160883 |
| Unspecified as acute or chronic, without haemorrhage or perforation | 1.11 (0.73,1.68) | 0.890 | 2423/159937 |
| Nerve root and plexus compressions in intervertebral disk disorders | 1.11 (0.72,1.71) | 0.890 | 2256/160104 |
| Faecal incontinence | 1.12 (0.71,1.77) | 0.890 | 2024/160336 |
| Pain localised to other parts of lower abdomen | 0.93 (0.68,1.26) | 0.890 | 4480/157880 |
| Hyperthyroidism/thyrotoxicosis* | 0.89 (0.57,1.41) | 0.890 | 2045/160295 |
| Haemorrhage of anus and rectum | 0.94 (0.72,1.22) | 0.890 | 6253/156107 |
| Precordial pain | 0.91 (0.61,1.35) | 0.890 | 2694/159666 |
| Headaches (not migraine)* | 1.14 (0.66,1.99) | 0.899 | 1360/160980 |
| Other senile cataract | 1.09 (0.76,1.56) | 0.905 | 3302/159058 |
| Polyuria | 0.89 (0.53,1.49) | 0.909 | 1575/160785 |
| Hypotension, unspecified | 1.08 (0.77,1.51) | 0.915 | 3807/158553 |
| Follow-up examination after other treatment for other conditions | 0.89 (0.52,1.51) | 0.916 | 1494/160866 |
| Arthrosis, unspecified (Ankle and foot) | 0.91 (0.58,1.42) | 0.916 | 2119/160241 |
| Ascending colon | 0.89 (0.54,1.48) | 0.916 | 1657/160703 |
| Migraine* | 1.06 (0.81,1.38) | 0.917 | 6315/156025 |
| Other specified cataract | 1.11 (0.68,1.82) | 0.917 | 1731/160629 |
| Epilepsy, unspecified | 0.90 (0.55,1.47) | 0.919 | 1792/160568 |
| Calculus of gallbladder with other cholecystitis | 0.93 (0.67,1.30) | 0.919 | 3948/158412 |
| Cough | 0.92 (0.61,1.38) | 0.919 | 2514/159846 |
| Hyperlipidaemia, unspecified | 1.08 (0.75,1.55) | 0.930 | 3226/159134 |
| Caecum | 1.11 (0.66,1.87) | 0.933 | 1538/160822 |
| Unspecified injury of head | 0.88 (0.46,1.67) | 0.934 | 1028/161332 |
| Unspecified diabates mellitus (Without complications) | 0.89 (0.49,1.61) | 0.939 | 1202/161158 |
| Surgical operation with implant of artificial internal device | 0.94 (0.68,1.30) | 0.939 | 4172/158188 |
| Other specified symptoms and signs involving the digestive system and abdomen | 1.11 (0.64,1.96) | 0.939 | 1323/161037 |
| Arthrosis of first carpometacarpal joint, unspecified | 1.13 (0.60,2.13) | 0.945 | 1029/161331 |
| Cystocele | 1.06 (0.78,1.43) | 0.953 | 4730/157630 |
| Disorders of calcium metabolism | 1.10 (0.64,1.89) | 0.962 | 1445/160915 |
| Cyst of kidney, acquired | 1.10 (0.60,2.02) | 0.977 | 1144/161216 |
| Follow-up examination after chemotherapy for other conditions | 0.90 (0.47,1.72) | 0.977 | 1004/161356 |
| Lumbar and other intervertebral disk disorders with radiculopathy | 1.07 (0.69,1.67) | 0.977 | 2191/160169 |
| Other specified abnormal uterine and vaginal bleeding | 0.90 (0.47,1.72) | 0.977 | 1008/161352 |
| Other specified diseases of liver | 1.10 (0.60,2.03) | 0.977 | 1119/161241 |
| Eye/eyelid problem* | 1.09 (0.64,1.84) | 0.977 | 1502/160838 |
| Unspecified lump in breast | 1.09 (0.63,1.89) | 0.977 | 1374/160986 |
| Personal history of diseases of the circulatory system | 1.03 (0.85,1.25) | 0.977 | 11966/150394 |
| Non-infective gastro-enteritis and colitis, unspecified | 1.04 (0.82,1.30) | 0.977 | 8495/153865 |
| Personal history of diseases of the nervous system and sense organs | 1.04 (0.81,1.34) | 0.977 | 6760/155600 |
| Other physical therapy | 1.04 (0.81,1.32) | 0.977 | 7586/154774 |
| Other and unspecified right bundle-branch block | 0.91 (0.49,1.68) | 0.977 | 1124/161236 |
| Heartburn | 1.08 (0.64,1.82) | 0.977 | 1555/160805 |
| Sciatica | 1.09 (0.61,1.93) | 0.977 | 1270/161090 |
| Cataract, unspecified | 0.97 (0.81,1.17) | 0.977 | 14230/148130 |
| Left ventricular failure | 0.94 (0.59,1.49) | 0.977 | 2003/160357 |
| Pain localised to upper abdomen | 1.03 (0.81,1.32) | 0.977 | 7125/155235 |
| Diverticular disease of large intestine without perforation or abscess | 1.02 (0.87,1.19) | 0.977 | 20324/142036 |
| Thyrotoxicosis, unspecified | 1.07 (0.63,1.81) | 0.977 | 1531/160829 |
| Postprocedural hypothyroidism | 1.07 (0.62,1.87) | 0.977 | 1366/160994 |
| Chronic sinusitis* | 0.92 (0.50,1.70) | 0.977 | 1135/161205 |
| Arthrosis, unspecified | 1.03 (0.84,1.26) | 0.977 | 10766/151594 |
| Duodenitis | 0.96 (0.68,1.35) | 0.977 | 3701/158659 |
| Personal history of other neoplasms | 0.96 (0.70,1.32) | 0.977 | 4206/158154 |
| Unspecified haemorrhoids without complication | 1.04 (0.78,1.38) | 0.977 | 5287/157073 |
| Occupational therapy and vocational rehabilitation, not elsewhere classified | 0.96 (0.70,1.32) | 0.977 | 4311/158049 |
| Calculus of bile duct without cholangitis or cholecystitis | 0.94 (0.59,1.52) | 0.977 | 1865/160495 |
| Alcohol use | 0.96 (0.70,1.32) | 0.977 | 4182/158178 |
| Personal history of infectious and parasitic diseases | 1.06 (0.67,1.66) | 0.977 | 2042/160318 |
| Presence of intraocular lens | 1.03 (0.81,1.30) | 0.977 | 8212/154148 |
| Localised oedema | 0.94 (0.57,1.57) | 0.977 | 1625/160735 |
| First degree haemorrhoids | 1.07 (0.58,1.98) | 0.977 | 1115/161245 |
| Hallux rigidus | 1.07 (0.59,1.94) | 0.977 | 1167/161193 |
| Polyarthrosis, unspecified | 0.97 (0.71,1.32) | 0.977 | 4564/157796 |
| Postmenopausal atrophic vaginitis | 0.95 (0.57,1.59) | 0.977 | 1580/160780 |
| Tonsiltis* | 1.06 (0.60,1.89) | 0.977 | 1256/161084 |
| Disorders of both mitral and tricuspid valves | 0.94 (0.50,1.77) | 0.977 | 1053/161307 |
| Glaucoma* | 0.95 (0.58,1.55) | 0.977 | 1773/160567 |
| Heart failure, unspecified | 1.05 (0.64,1.73) | 0.977 | 1699/160661 |
| Congestive heart failure | 1.05 (0.65,1.70) | 0.977 | 1830/160530 |
| Harmful use | 1.03 (0.79,1.34) | 0.977 | 6091/156269 |
| Spondylolisthesis (Lumbar region) | 1.07 (0.56,2.01) | 0.977 | 1037/161323 |
| Personal history of allergy to analgesic agent | 0.97 (0.72,1.31) | 0.977 | 4727/157633 |
| Intra-abdominal lymph nodes | 1.06 (0.57,1.97) | 0.977 | 1109/161251 |
| Other specified bacterial agents as the cause of diseases classified to other chapters | 0.96 (0.64,1.45) | 0.977 | 2524/159836 |
| Endometriosis of uterus | 1.06 (0.56,2.01) | 0.977 | 1035/161325 |
| Degeneration of macula and posterior pole | 1.03 (0.75,1.43) | 0.977 | 4074/158286 |
| Polyp of cervix uteri | 0.96 (0.65,1.42) | 0.977 | 2860/159500 |
| Allergy or anaphylactic reaction to drug* | 1.05 (0.64,1.72) | 0.977 | 1721/160619 |
| Pulmonary embolism +/- dvt* | 1.05 (0.60,1.84) | 0.977 | 1352/160988 |
| Other specified diseases of biliary tract | 0.94 (0.51,1.76) | 0.977 | 1088/161272 |
| Gastro-intestinal haemorrhage, unspecified | 1.03 (0.75,1.41) | 0.977 | 4340/158020 |
| Hypothyroidism/myxoedema* | 0.98 (0.82,1.18) | 0.977 | 13704/148636 |
| Mechanical complication of internal joint prosthesis | 1.05 (0.61,1.82) | 0.977 | 1384/160976 |
| Unspecified dementia | 0.95 (0.53,1.70) | 0.977 | 1265/161095 |
| Unspecified acute lower respiratory infection | 1.03 (0.77,1.36) | 0.977 | 5304/157056 |
| Rectocele | 1.03 (0.74,1.43) | 0.977 | 3880/158480 |
| Ear/vestibular disorder* | 0.95 (0.55,1.67) | 0.977 | 1361/160979 |
| Acute renal failure, unspecified | 0.98 (0.74,1.28) | 0.977 | 5897/156463 |
| Atrial fibrillation and atrial flutter, unspecified | 0.98 (0.75,1.28) | 0.977 | 6163/156197 |
| Breast cysts* | 1.05 (0.58,1.88) | 0.977 | 1218/161122 |
| Candidal stomatitis | 0.96 (0.52,1.76) | 0.977 | 1131/161229 |
| Other primary coxarthrosis | 1.03 (0.68,1.58) | 0.977 | 2366/159994 |
| Dysphagia | 1.02 (0.77,1.36) | 0.977 | 5349/157011 |
| Presence of other cardiac and vascular implants and grafts | 0.96 (0.50,1.81) | 0.977 | 1027/161333 |
| Family history of ischaemic heart disease and other diseases of the circulatory system | 0.98 (0.77,1.26) | 0.977 | 6974/155386 |
| Personal history of allergy to narcotic agent | 1.03 (0.67,1.59) | 0.977 | 2226/160134 |
| Actinic keratosis | 1.03 (0.65,1.65) | 0.977 | 1921/160439 |
| Malignant melanoma* | 1.04 (0.61,1.77) | 0.977 | 1466/160874 |
| Spinal stenosis (Lumbar region) | 0.97 (0.63,1.49) | 0.977 | 2325/160035 |
| Acquired absence of other parts of digestive tract | 1.02 (0.79,1.31) | 0.977 | 6749/155611 |
| Bronchiectasis | 1.03 (0.67,1.58) | 0.977 | 2272/160088 |
| Other specified disorders of bladder | 0.98 (0.67,1.43) | 0.977 | 2962/159398 |
| Old myocardial infarction | 0.98 (0.69,1.39) | 0.977 | 3459/158901 |
| Prolapsed disc/slipped disc* | 1.03 (0.67,1.56) | 0.977 | 2404/159936 |
| Other specified diseases of intestine | 0.97 (0.54,1.73) | 0.977 | 1243/161117 |
| Elevated blood-pressure reading, without diagnosis of hypertension | 1.04 (0.55,1.95) | 0.977 | 1059/161301 |
| Retention of urine | 1.02 (0.71,1.47) | 0.977 | 3259/159101 |
| Gastro-oesophageal reflux (gord) / gastric reflux* | 0.99 (0.77,1.26) | 0.979 | 7106/155234 |
| Oesophagitis | 1.02 (0.76,1.36) | 0.979 | 5076/157284 |
| Sigmoid colon | 0.98 (0.69,1.41) | 0.988 | 3329/159031 |
| Tachycardia, unspecified | 0.98 (0.62,1.54) | 0.988 | 2057/160303 |
| Barrett's oesophagus | 0.98 (0.60,1.60) | 0.988 | 1771/160589 |
| Procedure not carried out, unspecified reason | 0.98 (0.57,1.66) | 0.988 | 1489/160871 |
| Unspecified fall (Home) | 1.02 (0.64,1.62) | 0.992 | 1949/160411 |
| Chronic gastritis, unspecified | 0.98 (0.64,1.52) | 0.992 | 2283/160077 |
| Ventral hernia without obstruction or gangrene | 1.02 (0.59,1.76) | 0.992 | 1408/160952 |
| Anxiety/panic attacks* | 0.99 (0.66,1.49) | 0.992 | 2538/159802 |
| Other forms of chronic ischaemic heart disease | 0.98 (0.57,1.70) | 0.992 | 1423/160937 |
| Calculus of gallbladder with acute cholecystitis | 1.02 (0.56,1.85) | 0.992 | 1168/161192 |
| Iron deficiency anaemia, unspecified | 1.01 (0.76,1.33) | 0.992 | 5704/156656 |
| Personal history of diseases of the digestive system | 1.01 (0.82,1.23) | 0.992 | 11240/151120 |
| Excessive and frequent menstruation with irregular cycle | 0.99 (0.57,1.72) | 0.992 | 1367/160993 |
| Change in bowel habit | 0.99 (0.80,1.24) | 0.992 | 8943/153417 |
| Primary open-angle glaucoma | 0.99 (0.55,1.77) | 0.992 | 1228/161132 |
| Dorsalgia, unspecified | 0.99 (0.67,1.47) | 0.996 | 2744/159616 |
| Unilateral or unspecified inguinal hernia, without obstruction or gangrene | 0.99 (0.55,1.77) | 0.996 | 1250/161110 |
| Haemorrhage and haematoma complicating a procedure, not elsewhere classified | 1.01 (0.69,1.46) | 0.996 | 3054/159306 |
| Bradycardia, unspecified | 0.99 (0.60,1.64) | 0.996 | 1690/160670 |
| Arthritis, unspecified | 1.00 (0.78,1.29) | 0.996 | 6917/155443 |
| Leiomyoma of uterus, unspecified | 1.00 (0.75,1.34) | 1.000 | 5127/157233 |
| Derangement of meniscus due to old tear or injury (Medial collateral ligament or Other and unspecified medial meniscus) | 1.00 (0.58,1.72) | 1.000 | 1404/160956 |
| Cataract* | 1.00 (0.68,1.47) | 1.000 | 2886/159454 |
| Rheumatoid arthritis* | 1.00 (0.67,1.50) | 1.000 | 2620/159720 |
| Other specified abnormal findings of blood chemistry | 1.00 (0.68,1.47) | 1.000 | 2887/159473 |
| Tendency to fall, not elsewhere classified | 1.00 (0.73,1.38) | 1.000 | 4229/158131 |
| Residual haemorrhoidal skin tags | 1.00 (0.61,1.64) | 1.000 | 1707/160653 |
| Personal history of diseases of the genito-urinary system | 1.00 (0.76,1.32) | 1.000 | 5632/156728 |
| Other specified cerebrovascular diseases | 1.00 (0.60,1.66) | 1.000 | 1634/160726 |

*Self-reported outcomes recorded at UK Biobank recruitment interview.

The table presents outcomes included in the PheWAS, odds ratios scaled to reflect the effect per standard deviation decrease in genetically-proxied serum oestradiol with corresponding 95% confidence intervals, FDR-adjusted p-values, and the number of cases and controls for each outcome.
