## Supplementary Table 3 for "Genetic prediction of long-term effects of aromatase inhibition on cancer and non-neoplastic disease risk"

**Supplementary Table 3. Subgroup analyses results for rs727479 and population subgroups**

| Subgroup | Outcome | Cases | Effect size (95% CI) | P-value | P_LRT_ |
| --- | --- | --- | --- | --- | --- |
| BMI |  |  |  |  |  |
|  | Endometrial cancer | |  |  | 0.73 |
|  | Tertile 1 (≤ 25) | 483 | 0.91 (0.80,1.04) | 0.186 |  |
|  | Tertile 2 (> 25 & < 30) | 650 | 0.95 (0.85,1.07) | 0.400 |  |
|  | Tertile 3 (>30) | 310 | 0.98 (0.83,1.15) | 0.768 |  |
|  | Breast cancer | |  |  | 0.80 |
|  | Tertile 1 (≤ 25) | 5148 | 0.98 (0.94,1.02) | 0.353 |  |
|  | Tertile 2 (> 25 & < 30) | 5453 | 0.96 (0.92,1.00) | 0.042 |  |
|  | Tertile 3 (>30) | 3565 | 0.96 (0.91,1.01) | 0.095 |  |
|  | Polyps of corpus uteri | | |  | 0.001 |
|  | Tertile 1 (≤ 25) | 3025 | 1.02 (0.97,1.08) | 0.361 |  |
|  | Tertile 2 (> 25 & < 30) | 3354 | 0.93 (0.89,0.98) | 0.008 |  |
|  | Tertile 3 (>30) | 3043 | 0.87 (0.83,0.92) | < 0.001 |  |
|  | Postmenopausal bleeding | | |  | 0.02 |
|  | Tertile 1 (≤ 25) | 3762 | 0.97 (0.93,1.02) | 0.295 |  |
|  | Tertile 2 (> 25 & < 30) | 4065 | 0.96 (0.92,1.01) | 0.095 |  |
|  | Tertile 3 (>30) | 3409 | 0.90 (0.85,0.94) | < 0.001 |  |
|  | Chemotherapy session | | |  | 0.85 |
|  | Tertile 1 (≤ 25) | 2682 | 0.95 (0.90,1.01) | 0.108 |  |
|  | Tertile 2 (> 25 & < 30) | 2797 | 0.94 (0.89,1.00) | 0.042 |  |
|  | Tertile 3 (>30) | 1792 | 0.92 (0.85,0.98) | 0.014 |  |
|  | Distal radius fracture | | |  | 0.90 |
|  | Tertile 1 (≤ 25) | 2102 | 1.09 (1.02,1.16) | 0.010 |  |
|  | Tertile 2 (> 25 & < 30) | 1789 | 1.08 (1.01,1.16) | 0.020 |  |
|  | Tertile 3 (>30) | 822 | 1.10 (0.99,1.21) | 0.070 |  |
|  | Malaise and fatigue | |  |  | 0.79 |
|  | Tertile 1 (≤ 25) | 1293 | 0.92 (0.85,1.00) | 0.048 |  |
|  | Tertile 2 (> 25 & < 30) | 1362 | 0.94 (0.87,1.02) | 0.156 |  |
|  | Tertile 3 (>30) | 1070 | 0.98 (0.90,1.07) | 0.705 |  |
|  | Infection following procedure | | |  | 0.79 |
|  | Tertile 1 (≤ 25) | 861 | 0.92 (0.83,1.02) | 0.111 |  |
|  | Tertile 2 (> 25 & < 30) | 1081 | 0.88 (0.80,0.96) | 0.004 |  |
|  | Tertile 3 (>30) | 1268 | 0.97 (0.89,1.05) | 0.480 |  |
|  | Heel BMD* |  |  |  | 0.80 |
|  | Tertile 1 (≤ 25) | 48404 | -0.003 (-0.004,-0.001) | 0.001 |  |
|  | Tertile 2 (> 25 & < 30) | 45731 | -0.003 (-0.004,-0.001) | < 0.001 |  |
|  | Tertile 3 (>30) | 27944 | -0.003 (-0.005,-0.001) | 0.002 |  |
| Testosterone:SHBG | |  |  |  |  |
|  | Endometrial cancer | |  |  | 0.14 |
|  | Tertile 1 (0.00157 to 0.0133) | 310 | 0.98 (0.83,1.15) | 0.768 |  |
|  | Tertile 2 (0.01333 to 0.0238) | 430 | 1.02 (0.89,1.17) | 0.779 |  |
|  | Tertile 3 (0.02375 to 1.4633) | 751 | 0.80 (0.71,0.89) | 0.0001 |  |
|  | Breast cancer | |  |  | 0.79 |
|  | Tertile 1 (0.00157 to 0.0133) | 3208 | 0.98 (0.93,1.04) | 0.529 |  |
|  | Tertile 2 (0.01333 to 0.0238) | 3454 | 0.92 (0.88,0.97) | 0.002 |  |
|  | Tertile 3 (0.02375 to 1.4633) | 3733 | 0.99 (0.94,1.04) | 0.604 |  |
|  | Polyps of corpus uteri | | |  | 0.16 |
|  | Tertile 1 (0.00157 to 0.0133) | 1918 | 1.04 (0.97,1.11) | 0.284 |  |
|  | Tertile 2 (0.01333 to 0.0238) | 2219 | 0.98 (0.92,1.04) | 0.479 |  |
|  | Tertile 3 (0.02375 to 1.4633) | 2941 | 0.89 (0.84,0.94) | < 0.001 |  |
|  | Postmenopausal bleeding | | |  | 0.09 |
|  | Tertile 1 (0.00157 to 0.0133) | 2414 | 1.02 (0.96,1.08) | 0.599 |  |
|  | Tertile 2 (0.01333 to 0.0238) | 2490 | 0.98 (0.93,1.04) | 0.513 |  |
|  | Tertile 3 (0.02375 to 1.4633) | 3226 | 0.89 (0.85,0.94) | < 0.001 |  |
|  | Chemotherapy session | | |  | 0.87 |
|  | Tertile 1 (0.00157 to 0.0133) | 1705 | 0.93 (0.86,1.00) | 0.043 |  |
|  | Tertile 2 (0.01333 to 0.0238) | 1655 | 0.94 (0.87,1.01) | 0.077 |  |
|  | Tertile 3 (0.02375 to 1.4633) | 1729 | 0.96 (0.89,1.03) | 0.246 |  |
|  | Distal radius fracture | | |  | 0.79 |
|  | Tertile 1 (0.00157 to 0.0133) | 1322 | 1.10 (1.02,1.20) | 0.014 |  |
|  | Tertile 2 (0.01333 to 0.0238) | 1077 | 1.09 (1.00,1.19) | 0.053 |  |
|  | Tertile 3 (0.02375 to 1.4633) | 876 | 1.07 (0.97,1.18) | 0.163 |  |
|  | Malaise and fatigue | |  |  | 0.79 |
|  | Tertile 1 (0.00157 to 0.0133) | 830 | 1.00 (0.90,1.10) | 0.923 |  |
|  | Tertile 2 (0.01333 to 0.0238) | 851 | 0.94 (0.85,1.04) | 0.206 |  |
|  | Tertile 3 (0.02375 to 1.4633) | 815 | 0.95 (0.86,1.06) | 0.377 |  |
|  | Infection following procedure | | |  | 0.79 |
|  | Tertile 1 (0.00157 to 0.0133) | 658 | 0.85 (0.76,0.96) | 0.007 |  |
|  | Tertile 2 (0.01333 to 0.0238) | 709 | 0.97 (0.87,1.08) | 0.543 |  |
|  | Tertile 3 (0.02375 to 1.4633) | 875 | 0.96 (0.87,1.06) | 0.376 |  |
|  | Heel BMD* |  |  |  | 0.94 |
|  | Tertile 1 (0.00157 to 0.0133) | 29243 | -0.002 (-0.004,0.000) | 0.039 |  |
|  | Tertile 2 (0.01333 to 0.0238) | 29824 | -0.004 (-0.006,-0.002) | < 0.001 |  |
|  | Tertile 3 (0.02375 to 1.4633) | 29797 | -0.002 (-0.004,0.000) | 0.018 |  |
| BMD PRS | |  |  |  |  |
|  | Endometrial cancer | |  |  | 0.99 |
|  | Tertile 1 (-3.65e-06 to -3.11e-07) | 657 | 0.91 (0.81,1.02) | 0.117 |  |
|  | Tertile 2 (-3.11e-07 to 3.16e-07) | 687 | 0.86 (0.77,0.96) | 0.009 |  |
|  | Tertile 3 (3.16e-07 to 3.53e-06) | 685 | 0.91 (0.81,1.02) | 0.105 |  |
|  | Breast cancer | |  |  | 0.79 |
|  | Tertile 1 (-3.65e-06 to -3.11e-07) | 4687 | 0.97 (0.93,1.01) | 0.134 |  |
|  | Tertile 2 (-3.11e-07 to 3.16e-07) | 4800 | 0.97 (0.93,1.01) | 0.122 |  |
|  | Tertile 3 (3.16e-07 to 3.53e-06) | 4717 | 0.97 (0.93,1.01) | 0.118 |  |
|  | Polyps of corpus uteri | | |  | 0.66 |
|  | Tertile 1 (-3.65e-06 to -3.11e-07) | 3091 | 0.93 (0.88,0.98) | 0.004 |  |
|  | Tertile 2 (-3.11e-07 to 3.16e-07) | 3170 | 0.92 (0.87,0.97) | 0.002 |  |
|  | Tertile 3 (3.16e-07 to 3.53e-06) | 3198 | 0.97 (0.92,1.03) | 0.326 |  |
|  | Postmenopausal bleeding | | |  | 0.14 |
|  | Tertile 1 (-3.65e-06 to -3.11e-07) | 3730 | 0.92 (0.88,0.97) | 0.001 |  |
|  | Tertile 2 (-3.11e-07 to 3.16e-07) | 3854 | 0.94 (0.89,0.98) | 0.008 |  |
|  | Tertile 3 (3.16e-07 to 3.53e-06) | 3710 | 0.98 (0.93,1.03) | 0.341 |  |
|  | Chemotherapy session | | |  | 0.79 |
|  | Tertile 1 (-3.65e-06 to -3.11e-07) | 2509 | 0.94 (0.89,1.00) | 0.050 |  |
|  | Tertile 2 (-3.11e-07 to 3.16e-07) | 2451 | 0.94 (0.89,1.00) | 0.046 |  |
|  | Tertile 3 (3.16e-07 to 3.53e-06) | 2341 | 0.93 (0.88,0.99) | 0.026 |  |
|  | Distal radius fracture | | |  | 0.85 |
|  | Tertile 1 (-3.65e-06 to -3.11e-07) | 1655 | 1.06 (0.99,1.14) | 0.103 |  |
|  | Tertile 2 (-3.11e-07 to 3.16e-07) | 1608 | 1.13 (1.05,1.21) | 0.001 |  |
|  | Tertile 3 (3.16e-07 to 3.53e-06) | 1468 | 1.07 (0.99,1.15) | 0.087 |  |
|  | Malaise and fatigue | |  |  | 0.79 |
|  | Tertile 1 (-3.65e-06 to -3.11e-07) | 1201 | 0.95 (0.87,1.03) | 0.232 |  |
|  | Tertile 2 (-3.11e-07 to 3.16e-07) | 1230 | 0.93 (0.86,1.01) | 0.100 |  |
|  | Tertile 3 (3.16e-07 to 3.53e-06) | 1333 | 0.95 (0.88,1.03) | 0.238 |  |
|  | Infection following procedure | | |  | 0.87 |
|  | Tertile 1 (-3.65e-06 to -3.11e-07) | 1107 | 0.91 (0.83,0.99) | 0.036 |  |
|  | Tertile 2 (-3.11e-07 to 3.16e-07) | 1068 | 0.95 (0.87,1.04) | 0.265 |  |
|  | Tertile 3 (3.16e-07 to 3.53e-06) | 1061 | 0.91 (0.84,1.00) | 0.056 |  |
|  | Heel BMD* |  |  |  | 0.79 |
|  | Tertile 1 (-3.65e-06 to -3.11e-07) | 40493 | -0.002 (-0.004,-0.001) | 0.004 |  |
|  | Tertile 2 (-3.11e-07 to 3.16e-07) | 40971 | -0.003 (-0.004,-0.001) | 0.002 |  |
|  | Tertile 3 (3.16e-07 to 3.53e-06) | 40792 | -0.003 (-0.005,-0.002) | < 0.001 |  |
| Breast cancer PRS | |  |  |  |  |
|  | Endometrial cancer | |  |  | 0.87 |
|  | Tertile 1 (-1.78e-06 to -1.68e-07) | 629 | 0.87 (0.77,0.98) | 0.019 |  |
|  | Tertile 2 (-1.68e-07 to 1.66e-07) | 679 | 0.87 (0.78,0.98) | 0.017 |  |
|  | Tertile 3 (1.66e-07 to 1.81e-06) | 721 | 0.94 (0.84,1.05) | 0.274 |  |
|  | Breast cancer | |  |  | 0.85 |
|  | Tertile 1 (-1.78e-06 to -1.68e-07) | 3300 | 0.99 (0.94,1.04) | 0.710 |  |
|  | Tertile 2 (-1.68e-07 to 1.66e-07) | 4491 | 0.96 (0.92,1.00) | 0.049 |  |
|  | Tertile 3 (1.66e-07 to 1.81e-06) | 6413 | 0.96 (0.93,1.00) | 0.032 |  |
|  | Polyps of corpus uteri | | |  | 0.79 |
|  | Tertile 1 (-1.78e-06 to -1.68e-07) | 2979 | 0.98 (0.92,1.03) | 0.371 |  |
|  | Tertile 2 (-1.68e-07 to 1.66e-07) | 3144 | 0.92 (0.87,0.97) | 0.002 |  |
|  | Tertile 3 (1.66e-07 to 1.81e-06) | 3336 | 0.93 (0.88,0.97) | 0.003 |  |
|  | Postmenopausal bleeding | | |  | 0.80 |
|  | Tertile 1 (-1.78e-06 to -1.68e-07) | 3671 | 0.96 (0.92,1.01) | 0.142 |  |
|  | Tertile 2 (-1.68e-07 to 1.66e-07) | 3724 | 0.92 (0.88,0.97) | 0.001 |  |
|  | Tertile 3 (1.66e-07 to 1.81e-06) | 3899 | 0.95 (0.91,0.99) | 0.030 |  |
|  | Chemotherapy session | | |  | 0.99 |
|  | Tertile 1 (-1.78e-06 to -1.68e-07) | 2086 | 0.96 (0.90,1.03) | 0.275 |  |
|  | Tertile 2 (-1.68e-07 to 1.66e-07) | 2086 | 0.96 (0.90,1.03) | 0.275 |  |
|  | Tertile 3 (1.66e-07 to 1.81e-06) | 2867 | 0.94 (0.89,0.99) | 0.018 |  |
|  | Distal radius fracture | | |  | 0.79 |
|  | Tertile 1 (-1.78e-06 to -1.68e-07) | 1578 | 1.07 (0.99,1.15) | 0.086 |  |
|  | Tertile 2 (-1.68e-07 to 1.66e-07) | 1532 | 1.08 (1.00,1.16) | 0.041 |  |
|  | Tertile 3 (1.66e-07 to 1.81e-06) | 1621 | 1.11 (1.04,1.20) | 0.003 |  |
|  | Malaise and fatigue | |  |  | 0.79 |
|  | Tertile 1 (-1.78e-06 to -1.68e-07) | 1240 | 0.90 (0.83,0.98) | 0.020 |  |
|  | Tertile 2 (-1.68e-07 to 1.66e-07) | 1262 | 0.95 (0.88,1.04) | 0.254 |  |
|  | Tertile 3 (1.66e-07 to 1.81e-06) | 1262 | 0.97 (0.90,1.06) | 0.539 |  |
|  | Infection following procedure | | |  | 0.79 |
|  | Tertile 1 (-1.78e-06 to -1.68e-07) | 1031 | 0.96 (0.88,1.06) | 0.440 |  |
|  | Tertile 2 (-1.68e-07 to 1.66e-07) | 1090 | 0.89 (0.81,0.97) | 0.008 |  |
|  | Tertile 3 (1.66e-07 to 1.81e-06) | 1115 | 0.93 (0.85,1.01) | 0.093 |  |
|  | Heel BMD* |  |  |  | 0.73 |
|  | Tertile 1 (-1.78e-06 to -1.68e-07) | 40501 | -0.002 (-0.004,-0.001) | 0.008 |  |
|  | Tertile 2 (-1.68e-07 to 1.66e-07) | 40907 | -0.003 (-0.005,-0.002) | < 0.001 |  |
|  | Tertile 3 (1.66e-07 to 1.81e-06) | 40848 | -0.003 (-0.005,-0.002) | < 0.001 |  |

The table presents hazard ratios (HRs) and corresponding 95% CIs for outcomes across tertiles of subgroup variables, p-values for the effect estimates, and the P_LRT_ for interaction between subgroup variable and outcome. HRs are scaled to reflect the effect per SD decrease in genetically-proxied serum oestradiol.

*Continuous outcome; effect size is expressed as SD change in the outcome per SD decrease in genetically-proxied serum oestradiol.
